# Source-level EEG and graph theory reveal widespread functional network alterations in focal epilepsy

**DOI:** 10.1101/2020.12.17.20248426

**Authors:** Christoffer Hatlestad-Hall, Ricardo Bruña, Marte Roa Syvertsen, Aksel Erichsen, Vebjørn Andersson, Fabrizio Vecchio, Francesca Miraglia, Paolo M. Rossini, Hanna Renvall, Erik Taubøll, Fernando Maestú, Ira H. Haraldsen

## Abstract

**Objective:** The hypersynchronous neuronal activity associated with epilepsy causes widespread functional network disruptions extending beyond the epileptogenic zone. This altered functional network topology is considered a mediator from which non-seizure symptoms arise, such as cognitive impairment. The aim of the present study was to demonstrate the presence of functional network alterations in focal epilepsy patients with good seizure control and high quality of life.

**Methods:** We compared twenty-two focal epilepsy patients and sixteen healthy controls on graph metrics derived from functional connectivity (phase-locking value) of source reconstructed resting-state EEG. Graph metrics were calculated over a predefined range of network densities in five frequency bands.

**Results:** In terms of global network topology alterations, we observed a significantly increased small world index in epilepsy patients relative to the healthy controls. On the local level, two left-hemisphere regions displayed a shift towards greater alpha band “hubness”.

**Conclusions:** Subtle widespread functional network alterations are evident in focal epilepsy, even in a cohort characterised by successful anti-seizure medication therapy and high quality of life. These findings suggest a possible clinical relevance of functional network analysis in epilepsy.

**Significance:** Focal epilepsy is accompanied by global and local functional network aberrancies which might be implied in the sustenance of non-seizure symptoms.

**Highlights:**

- Focal epilepsies are associated with widespread interictal functional network alterations, extending beyond the epilepsy focus.
- Global and local graph theoretical analyses of source-space EEG functional connectivity networks capture these network changes, and might thus be of clinical relevance.
- Group-level differences in network metrics are relatively stable across network analysis parameters.

## 1 Introduction

The human brain is a complex system which relies on coordinated and flexible network activity involving its constituent regions to regulate and sustain physiological processes, reaching from movement (King et al. 2018) and autonomic functions (Fan et al. 2012) to advanced cognition (Shine et al. 2019). Disruption of such networks is increasingly considered an important mediator of physiological dysfunction (Uhlhaas and Singer 2006). Thus, an aberrant network model may account for symptoms occurring in the absence of observable structural brain pathology. Characteristic in this respect is cognitive impairment, such as memory issues and executive dysfunctions (Holmes 2015).

The paragon of brain network pathology is epilepsy (Kramer and Cash 2012), which encompasses syndromes that are heterogeneous in aetiology and symptomatology, but share the clinical hallmark of acute transient disruption of brain function caused by hypersynchronous neuronal activity. These physiological processes often manifest as epileptic seizures, but may cause additional widespread cognitive dysfunction also present in interictal periods (van Diessen et al. 2013). In primarily focal epilepsies, the epileptic activity putatively originates in a limited pathological area (*i*.*e*., the epileptogenic zone) and may rapidly propagate to other cortical regions. Importantly, from a network perspective, local aberrancy may impact the organisation of networks at a larger scale, potentially causing subtle symptoms which are not directly associated with the classical functions of the epileptogenic brain area (Tailby et al. 2018). Identification and monitoring of these functional network changes in the brain may provide important clinical biomarkers for treatment of the wider range of symptoms associated with epileptic disorders (Haneef and Chiang 2014; Hallett et al. 2020).

In recent years, the methodological frameworks of functional brain connectivity (Bastos and Schoffelen 2015) and graph analysis (Behrens and Sporns 2012) have been increasingly used to characterise networks in the human brain. Functional connectivity (FC) is concerned with the statistical dependence between spatially separated signals obtained from electro- and magnetoencephalography (EEG/MEG) or functional magnetic resonance imaging (fMRI), and may be estimated on the basis of features related to the signals, such as amplitude, frequency and phase (van Diessen et al. 2015; Rossini et al. 2019).

From a clinical perspective, evidence suggests that network characteristics are capable of detecting differences between pathological conditions and healthy brains (van Straaten and Stam 2013; Olde Dubbelink et al. 2014; Stam 2014; Engels et al. 2015). In epilepsy, interictal network abnormalities have been consistently reported across imaging modalities, both in terms of alterations compared to healthy peers (Quraan et al. 2013; Niso et al. 2015; Výtvarová et al. 2017) and as a biomarker for cognitive status (Vlooswijk et al. 2011; Rodríguez-Cruces et al. 2020). However, the clinical use of FC and graph analysis is arguably still in a rudimentary stage (Douw et al. 2019), as methodology and findings diverge substantially across studies (Vlooswijk et al. 2011; Quraan et al. 2013; Pedersen et al. 2015), and there remain unresolved issues concerning analysis parameters, such as network threshold scheme (van den Heuvel et al. 2010; van Wijk et al. 2010; Garrison et al. 2015) and underlying connectivity measures (Horstmann et al. 2010; Niso et al. 2015).

Nevertheless, the most prominent of these issues with regard to clinical epileptology concerns the choice of functional imaging modality. Currently, investigations targeting connectivity and network analysis in epilepsy are dominated by fMRI (Vlooswijk et al. 2011; McCormick et al. 2013; Vaughan et al. 2016; Výtvarová et al. 2017). However, the rapidly evolving dynamics of brain networks might be better captured by the millisecond timescale of MEG (Elshahabi et al. 2015; Niso et al. 2015) and EEG (Quraan et al. 2013; Vecchio et al. 2015). Importantly, the electrophysiological approach allows investigations into oscillatory activity in specific frequency bands (Rossini et al. 2019), and by extension, their associated cognitive functions (for a review, see Lopes da Silva 2013). Despite its arguably superior signal-to-noise ratio, MEG remains scarcely available in clinical settings worldwide, and the position of EEG in epilepsy diagnosis is pervasive. However, graph theory studies of focal epilepsy via EEG-based functional networks are few, and sample sizes are generally limited (Horstmann et al. 2010; Quraan et al. 2013; Vecchio et al. 2015). Thus, to achieve a feasible means of utilising network measures in applied neurology, EEG-based network methodology warrants development.

Important in this regard is to mitigate the impact of noise and volume conduction on scalp-recorded EEG (Brunner et al. 2016), which constitute an ubiquitous, non-specific perturbation of connectivity estimates. Methods of source reconstruction have been developed (Schoffelen and Gross 2009; Vorwerk et al. 2014; Hassan and Wendling 2018), which have been shown to reduce these effects in EEG (Besserve et al. 2011). Provided further rigorous methodological assessment and development, EEG/MEG source-space connectivity is envisioned a future central role in both diagnostics and treatment of epilepsy (van Mierlo et al. 2019).

The aim of the present study was to examine brain network organisation in a cohort of focal epilepsy patients with good seizure control and high quality of life, and age-matched healthy control subjects. The investigated networks were constructed with bivariate estimates of EEG-based source-space phase-locking value (PLV), under the hypothesis of phase synchronisation; *i*.*e*., functionally connected brain regions generate signals whose phases evolve together (Rosenblum et al. 1996). To assay the stability of our findings, we evaluated the global and local network metrics under a range of predefined network densities. Provided the well-functioning epilepsy patients in the study, our hypothesis was that only subtle interictal network alterations would be observed compared to non-epileptic subjects. Importantly, it was not within the scope of this study to investigate clinical factors relating to the individual patient, but rather to assess group-level network alterations in a heterogenous focal epilepsy cohort. This exploratory effort aims to contribute to the following issues: (1) corroborate previous reports of alterations in global functional network organisation in focal epilepsy; (2) aid the use and interpretation of macro level local graph metrics in focal epilepsy; and (3) evaluate the effect of network matrix threshold parameters for the stability of group differences in clinical graph analysis application.

## 2 Materials and methods

### 2.1 Participants

Twenty-two patients diagnosed with uni- or bilateral focal epilepsy (FE; fourteen females, age 55.1 ± 4.3 years) were recruited from neurological outpatient clinics, in connection with routine follow-up visits. All patients were characterised by high levels of daily functioning. In addition, sixteen age-matched healthy control (HC) subjects (twelve females, age 55.9 ± 6.9 years) participated. The HC participants were recruited from the FE patients’ social networks, providing controls with similar socio-economic background to the patients. All FE subjects had used the same anti-seizure medication (ASM) for at least six months prior to their participation in the study. Neither FE nor HC subjects had any history of epilepsy surgery, psychiatric disorders, developmental disorders, or any other debilitating diseases. The patients’ clinical data, including ASM therapy, aetiology, EEG/MRI pathology and focus localisation, are listed patient-wise in Table 1. Of the eight patients with MRI-verified pathology, two had mesial temporal sclerosis, whereas the remaining six had other local structural pathology. None had indications of progressive neurological disease nor tumour. All patients presented with past or present focal seizures. Twelve of whom also had focal seizures with secondary generalisation. Neither FE nor HC participants were compensated for their study participation. Ethical approval for the study was granted by the Regional Ethics Committee of South-Eastern Norway. All participants provided informed written consent in accordance with the Declaration of Helsinki.

**Table 1:**
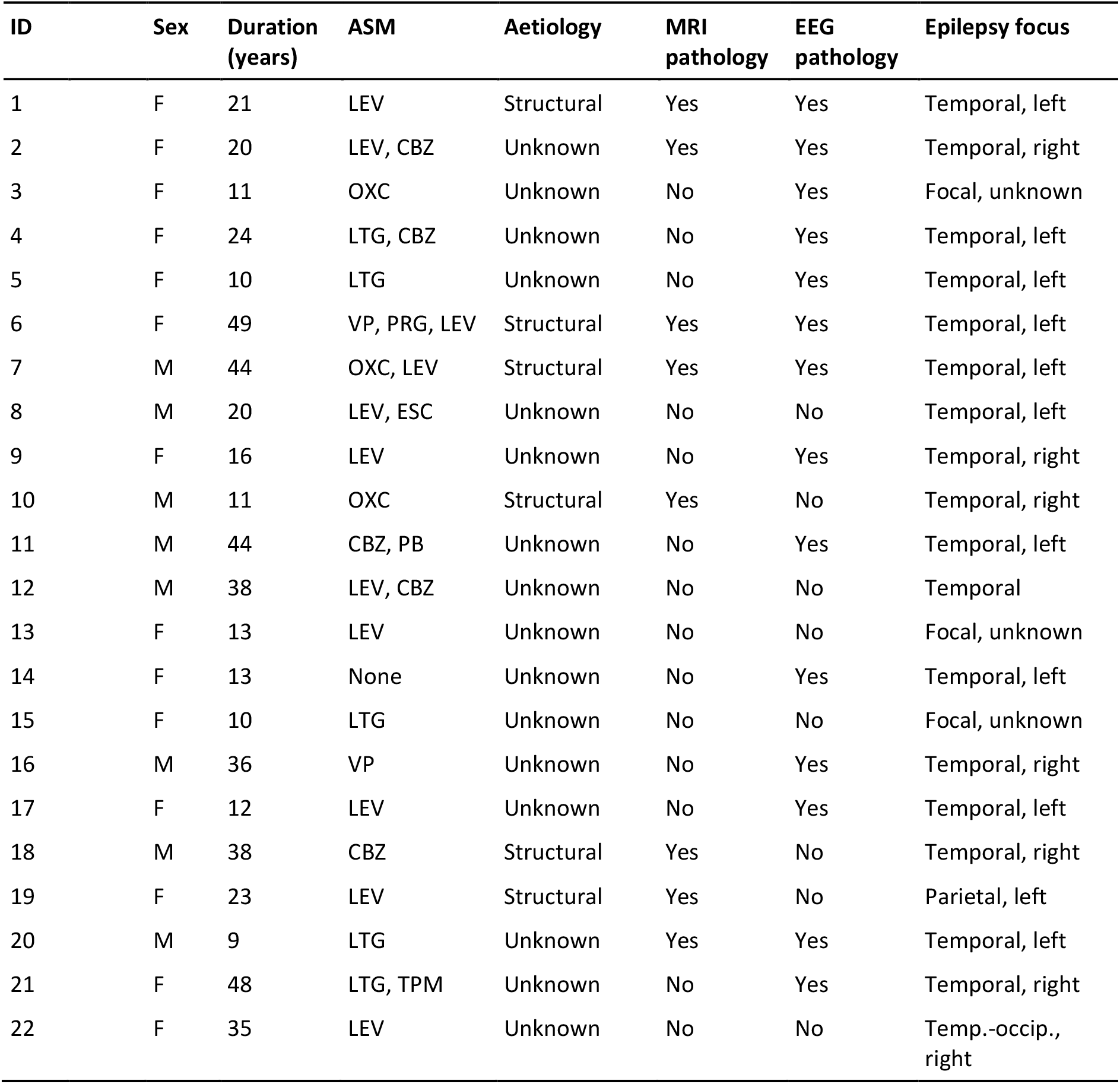
Clinical data of the patients. Sex: F = female; M = male. Duration of epilepsy is defined as years since the patient’s first seizure. Anti-seizure medications (ASM): CBZ = Carbamazepine; OXC = Oxcarbazepine; ESC = Eslicarbazepine acetate; LEV = Levetiracetam; LTG = Lamotrigine; PRG = Pregabalin; TPM = Topiramate; VP = Valproate. The MRI/EEG pathology columns denote whether the patient presents with pathological findings. Epilepsy focus reflects the clinical diagnosis made on the basis of all available information.

| ID | Sex | Duration (years) | ASM | Aetiology | MRI pathology | EEG pathology | Epilepsy focus |
| --- | --- | --- | --- | --- | --- | --- | --- |
| 1 | F | 21 | LEV | Structural | Yes | Yes | Temporal, left |
| 2 | F | 20 | LEV, CBZ | Unknown | Yes | Yes | Temporal, right |
| 3 | F | 11 | OXC | Unknown | No | Yes | Focal, unknown |
| 4 | F | 24 | LTG, CBZ | Unknown | No | Yes | Temporal, left |
| 5 | F | 10 | LTG | Unknown | No | Yes | Temporal, left |
| 6 | F | 49 | VP, PRG, LEV | Structural | Yes | Yes | Temporal, left |
| 7 | M | 44 | OXC, LEV | Structural | Yes | Yes | Temporal, left |
| 8 | M | 20 | LEV, ESC | Unknown | No | No | Temporal, left |
| 9 | F | 16 | LEV | Unknown | No | Yes | Temporal, right |
| 10 | M | 11 | OXC | Structural | Yes | No | Temporal, right |
| 11 | M | 44 | CBZ, PB | Unknown | No | Yes | Temporal, left |
| 12 | M | 38 | LEV, CBZ | Unknown | No | No | Temporal |
| 13 | F | 13 | LEV | Unknown | No | No | Focal, unknown |
| 14 | F | 13 | None | Unknown | No | Yes | Temporal, left |
| 15 | F | 10 | LTG | Unknown | No | No | Focal, unknown |
| 16 | M | 36 | VP | Unknown | No | Yes | Temporal, right |
| 17 | F | 12 | LEV | Unknown | No | Yes | Temporal, left |
| 18 | M | 38 | CBZ | Structural | Yes | No | Temporal, right |
| 19 | F | 23 | LEV | Structural | Yes | No | Parietal, left |
| 20 | M | 9 | LTG | Unknown | Yes | Yes | Temporal, left |
| 21 | F | 48 | LTG, TPM | Unknown | No | Yes | Temporal, right |
| 22 | F | 35 | LEV | Unknown | No | No | Temp.-occip., right |

### 2.2 EEG acquisition and preprocessing

EEG was recorded with a BioSemi 128-channel system (sampling rate of 2048 Hz) during a resting-state period. For this segment, the participant was comfortably seated in a chair, resting with his or her eyes closed, but awake, for 4 minutes. To minimise between-subject variation regarding task comprehension, instructions were given in written form on a screen, as recommended by van Diessen and colleagues (2015).

High-precision information on the spatial locations of the EEG electrodes was acquired using an IO Structure Sensor (Occipital, Inc.) scanner device for iPad (Apple, Inc.). From the 3D model, the electrodes were spatially identified by an operator using MATLAB (The MathWorks, Inc.) and FieldTrip functions (ver. 2019-01-16; Oostenveld et al. 2011). For enhanced accuracy in the manual alignment of electrode positions and head model (see below), the head shape mesh was also extracted.

The EEG data were preprocessed using EEGLAB functions (ver. 2019.1; Delorme and Makeig 2004). The signals were downsampled to 512 Hz, and re-referenced to an average reference, obtained by iteratively removing noisy signals (amplitude SD larger than 25 µV) from the reference signal composite. Segments containing high-amplitude signals (amplitude larger than 150 µV in more than 25% of the signals) were removed, and consistently noisy signals were removed with the PREP Pipeline toolbox (Bigdely-Shamlo et al. 2015). Line noise (50 Hz and harmonics) was suppressed with Zapline (de Cheveigné 2020). To correct artefacts related to ocular and muscular activity, the signals were decomposed with the Second Order Blind Identification (SOBI; Belouchrani et al. 1993) algorithm, and categorised with ICLabel (Pion-Tonachini et al. 2019). The signals were band-pass filtered between 1 and 45 Hz (EEGLAB default settings). An additional 10 seconds of mirrored data was temporarily applied at both onset and offset to avoid edge artifacts. Finally, the data were segmented into non-overlapping epochs of 4 seconds, and visually evaluated. Noise- and artefact-free epochs were included for source reconstruction.

### 2.3 EEG source reconstruction and functional connectivity

From each participant, a T1-weighted MRI image was acquired using either one of two scanners: a 3T Philips Achieva or a 1.5T Siemens Magnetom Aera. This anatomical image was segmented using the SPM12 Unified Segmentation algorithm (Ashburner and Friston 2005) and the NY Head tissue probability map (Huang et al. 2016) to create a three-layer boundary element conduction model (BEM). The surfaces were generated using the iso2mesh toolbox (Qianqian Fang and Boas 2009) for MATLAB.

The source model consisted of a homogeneous regular grid of dipoles defined in MNI space, with a uniform separation of 10 mm, and labeled according to the Automated Anatomical Labeling (AAL; Tzourio-Mazoyer et al. 2002) atlas. The final grid consisted of 1210 dipoles belonging to one of the 80 cortical areas in the AAL atlas. This grid was linearly transformed to subject-space using the MRI, obtaining an individual source model. We combined this source model, the BEM conduction model and the individual electrode positions using OpenMEEG (Gramfort et al. 2010), obtaining an individual lead field. For nine subjects (five patients and four controls; all female) we were unable to obtain the individual head shape and electrode positions, and we used a set of electrode positions based on the average of the rest of participants.

As the inverse method, we used a spatial filter based on linearly constrained, minimum variance beamformers (Van Veen et al. 1997). The data used to build this spatial filter was band-pass filtered between 2 and 45 Hz by means of a FIR filter applied using 2 additional seconds of real data at each side as temporary padding.

The source-space FC was evaluated under the hypothesis of phase synchronisation (Rosenblum et al. 1996) using PLV (Lachaux et al. 1999; Bruña et al. 2018), which was estimated between pairs of 26 composite regions based on the AAL atlas (listed in Table 2; for spatial layout, see Fig. 1). This was done by calculating the root-mean-square of the PLV for all the links connecting one source position in one area with one source position in the other. Calculations were done separately for five frequency bands: theta (4-8 Hz), alpha (8-12 Hz), low-beta (12-20 Hz), high-beta (20-30 Hz) and gamma (30-45 Hz).

**Table 2:** List of composite regions based on areas defined in the AAL atlas. The AAL area indices correspond to the original area/index pairing in Tzourio-Mazoyer et al. (2002). The region indices correspond to the map in Fig. 1.

| **Region index**<br>*Left/right* | **Region label** | **Composite of AAL areas**<br>*Left/right* |
| --- | --- | --- |
| 1/2 | Central region | 1/2, 17/18, 57/58 |
| 3/4 | Frontal lateral surface | 3/4, 7/8, 11/12, 13/14 |
| 5/6 | Frontal medial surface | 19/20, 23/24, 69/70 |
| 7/8 | Frontal orbital surface | 5/6, 9/10, 15/16, 21/22, 25/26, 27/28 |
| 9/10 | Temporal lateral surface | 79/80, 81/82, 85/86, 89/90 |
| 11/12 | Temporal medial surface (limbic) | 83/84, 87/88 |
| 13/14 | Parietal lateral surface | 59/60, 61/62, 63/64, 65/66 |
| 15/16 | Precuneus | 67/68 |
| 17/18 | Occipital lateral surface | 49/50, 51/52, 53/54 |
| 19/20 | Occipital medial and inferior surfaces | 43/44, 45/46, 47/48, 55/56 |
| 21/22 | Cingulum | 31/32, 33/34, 35/36 |
| 23/24 | Hippocampus | 37/38, 39/40 |
| 25/26 | Insula | 29/30 |

**Fig. 1:**
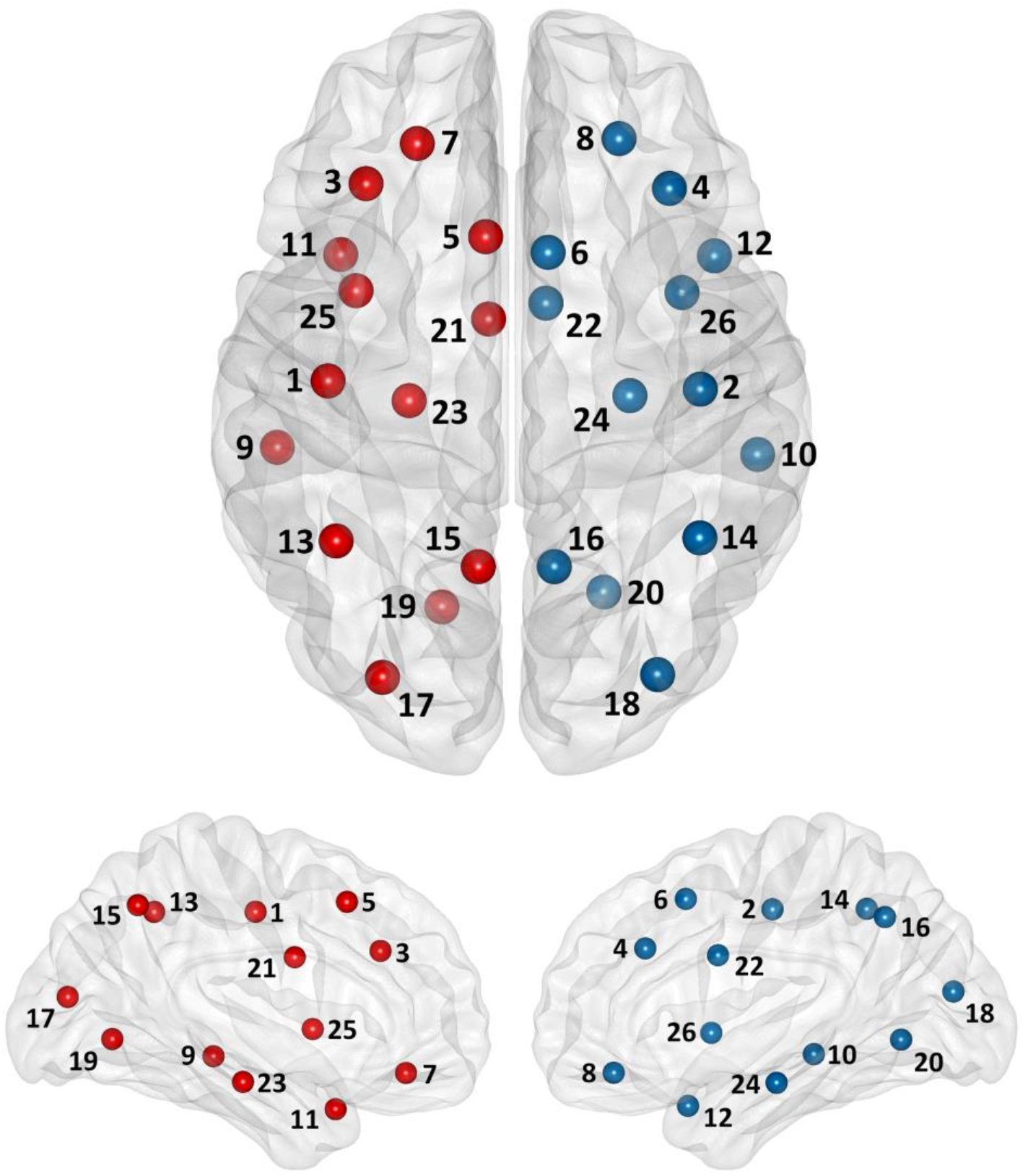
Node/brain region mapping. Node indices correspond to region indices listed in Table 2. Left hemisphere nodes are represented by red dots, whereas right hemisphere nodes are blue. The node positions are calculated from the centroid of the regions comprising the node. *Top*: Axial view; *bottom:* medial view.

### 2.4 Network analysis

The network analysis was conducted with functions from the Brain Connectivity Toolbox (BCT; ver. 2019-03-03; Rubinov and Sporns 2010) and in-house MATLAB code (available upon request). Each frequency band was analysed separately. Analyses were conducted on a range of predefined density values (López-Sanz et al. 2017; Sion et al. 2020). Graph metrics were calculated on both the whole-network (global) and the node (local) levels. In terms of network analysis, the FC matrices were *weighted* (PLV) and *undirected*.

All matrices were constructed with the constraint of being fully connected across all thresholds (*i*.*e*., no node was disconnected from the main network component). Full connectedness is a formal requirement in the definition of several global graph metrics. To implement this, we first computed the minimum spanning tree (MST; Stam et al. 2014; Tewarie et al. 2015) of the inverse FC matrix (formally, the maximum spanning tree of the FC matrix), and then added weights to the MST backbone incrementally by descending weight order until the density threshold was reached. The predefined density threshold (DT) range extended from 25 to 75%, with increments of 5%.

For each FC matrix, one hundred re-wired null models with preserved weight, degree and strength distributions were generated from the dense matrix (BCT: null_model_und_sign; Rubinov and Sporns 2011). These null models were processed identical to the empirical network, and the mean global network metrics calculated from them were used to normalise the corresponding metric for the empirical network.

The following graph metrics were calculated: The *clustering coefficient*, a quantification of the density of connections between a node’s neighbours. The *characteristic path length*, which describes the network’s tendency towards global integration and efficiency. The ratio between the normalised values of a network’s clustering coefficient and characteristic path length is often referred to as the *small world index*, a measure of to what degree the network architecture is balanced between local segregation and global integration (Watts and Strogatz 1998; Humphries and Gurney 2008). The *node strength*, defined as the sum of all edges connecting a node to other nodes. Finally, the *eigenvector centrality*, which is a measure of a node’s relative importance to the network; it takes into account the number and strength of the node’s edges, and whether these edges connect the node to other central nodes. For comprehensive accounts of graph metrics and their mathematical definitions, the reader is referred to Rubinov & Sporns (2010) and Newman (2008).

### 2.5 Power spectra analysis

PLV has been observed to be reliable in test-retest scenarios (Colclough et al. 2016; Garcés et al. 2016). However, it has some caveats, the most important being its sensitivity to volume conduction and source leakage. Here, the former was mitigated by calculating PLV in source-space. However, it is impossible to remove the effects of source leakage. Individual differences in leakage may lead to differences in the magnitude of FC (Schoffelen and Gross 2009). The varying leakage is usually generated by differences in power in the involved regions (Muthukumaraswamy and Singh 2011). To account for this possibility, we conducted comparisons of frequency band-specific relative power spectra between groups. In the regions displaying stable local graph metric differences, we complemented the results with *post hoc* comparisons of power spectra.

### 2.6 Overall functional connectivity

Differences in the overall level of FC can spuriously generate differences in network metrics (van den Heuvel et al. 2017). In this context, overall FC is defined as the mean PLV in the individual FC matrix, discarding intra-regional estimates, and calculated separately for each frequency band. We explored possible differences in overall FC prior to conducting the analyses based on network metrics. These did not differ between groups.

### 2.7 Statistical tests

Taking into account the current study’s exploratory approach with relatively small samples, group differences in graph metrics on both global and local levels were tested with nonparametric permutation tests (Maris and Oostenveld 2007). For each comparison, 5000 permutations were carried out. The obtained *p* values were corrected for multiple comparisons across frequency bands (*i*.*e*., five comparisons) with the Benjamini-Yekutieli false discovery rate with assumed positive test correlation (FDR+) procedure (Benjamini and Yekutieli 2001; Genovese et al. 2002). FDR adjusted *p* values (*i*.*e*., *q* values) below 0.1 (Niso et al. 2015) were considered statistically significant. Uncorrected *p* values are reported in figures. Group differences were quantified as the corrected standardised mean difference, with the effect size estimate termed *Hedges’ g* (Lakens 2013).

Directionality of effects is consistently given such that positive values of *g* indicate larger values for the FE group than for the HC group. All comparisons were conducted independently for each DT level. For local graph metrics, findings were considered “stable” if significant frequency band-specific group differences were observed for a particular node in three or more consecutive DT increments. Mean *q* and *g* values were calculated across the significant DT levels of these node differences.

Group differences in overall FC and power spectra were analysed in an identical permutation procedure. In contrast to the exploratory analyses, the resulting *p* values from the overall FC tests were not corrected for multiple comparisons. This also applied to the relative power spectra comparison which was defined as a *post hoc* procedure for complementing the frequency band/node combinations which yielded significant group differences in local graph metrics. All statistical analyses were conducted with MATLAB.

## 3 Results

### 3.1 Global graph metrics

On the global level, group differences in normalised clustering coefficient and characteristic path length, and the small world index, were analysed. In line with our hypothesis, we observed subtle, yet consistent differences between patients and controls.

The networks of both patient and control groups displayed evidence of small world architecture (index larger than 1), which was represented in all frequency bands and across the majority of DT levels (Fig. 2, top row). However, in the upper areas of the DT range, the small world index fell below 1, suggesting that higher-density networks failed to display this global organisation. Upon comparison between the groups, the patients consistently showed higher indices, although at some DT levels, the direction of the difference was reversed (Fig. 2, bottom row). The group effects were most pronounced for the DT range between 60 and 70%, achieving statistical significance (Fig. 2, middle row) in the theta (DT = 65%: *q* = 0.074, *g* = 0.730), alpha (DT = 65%: *q* = 0.096, *g* = 0.629), high-beta (DT = 70%: *q* = 0.089, *g* = 0.706) and gamma bands (DT = 65/70%: *q* = 0.074/0.089, *g* = 0.766/0.714).

**Fig. 2:**
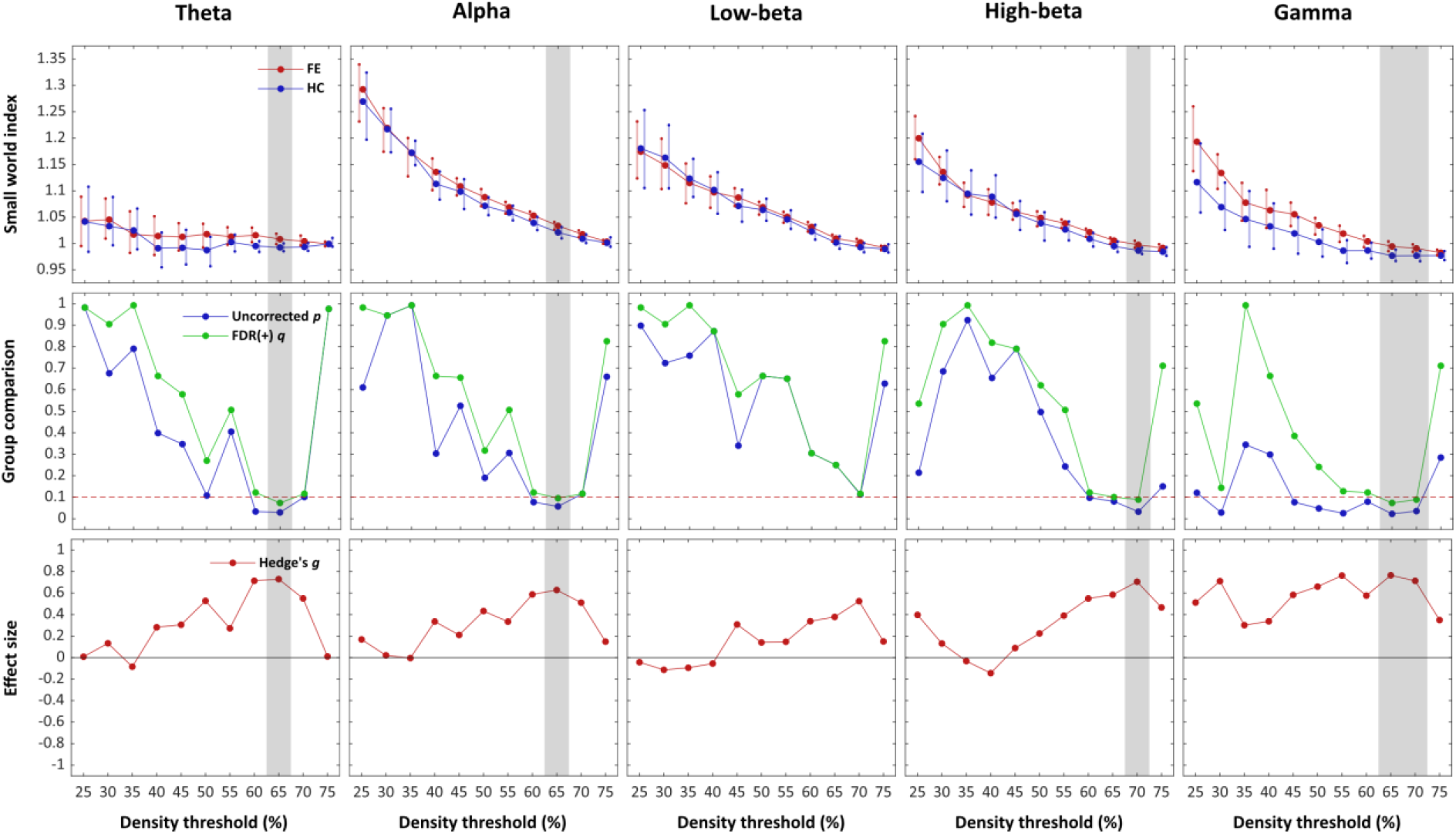
Small world index. Group comparison of the small world index across DT levels. The grey shadings indicate significant group difference at given DT level. *Top row:* Small world index, where the vertical lines represent the 95% BCa confidence interval of the mean. *Middle row:* Uncorrected *p* values and FDR adjusted *q* values associated with the permutation tests. The horizontal dashed line represents the critical alpha threshold (*q* < 0.1). *Bottom row:* Effect size estimates.

In addition, we analysed the normalised clustering coefficient (Fig. 3) and the normalised characteristic path length (Fig. 4) separately. Both patient and control groups displayed normalised values above 1, indicating that the observed network metrics were higher than the corresponding metrics in the randomly re-wired networks, in line with our expectations. The patients displayed consistently higher clustering coefficient and lower characteristic path length than the controls, but the effects did not reach statistical significance after FDR adjustment. Yet, these group differences showed evidence of stability across all frequency bands and DT levels, with the differences being the smallest in the low-beta band and most pronounced in the gamma band (clustering coefficient) and high-beta band (characteristic path length).

**Fig. 3:**
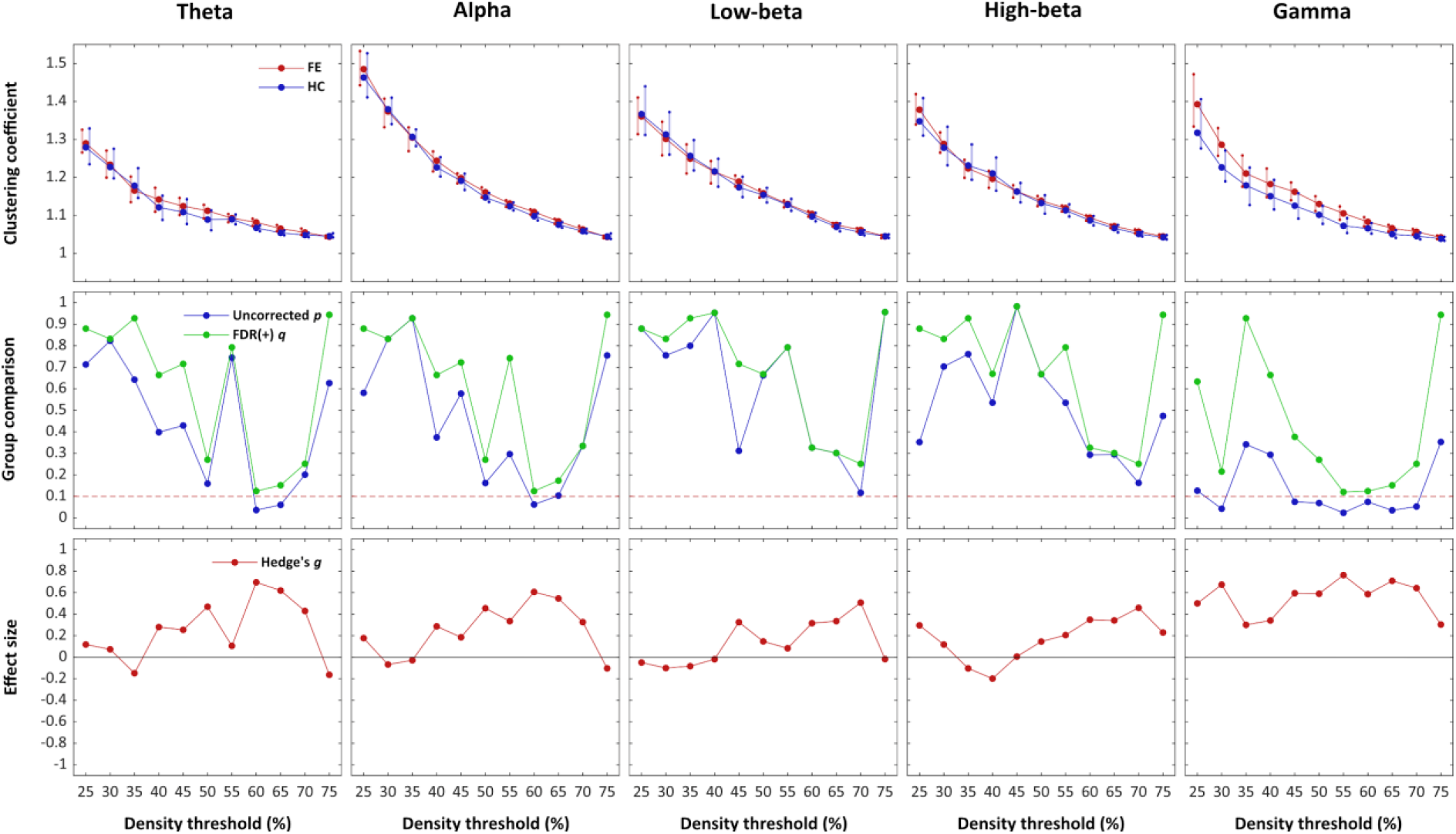
Normalised clustering coefficient. Group comparison of the network-wide normalised clustering coefficient across DT levels. *Top row:* Normalised clustering coefficient, where the vertical lines represent the 95% BCa confidence interval of the mean. *Middle row:* Uncorrected *p* values and FDR adjusted *q* values associated with the permutation tests. The horizontal dashed line represents the critical alpha threshold (*q* < 0.1). *Bottom row:* Effect size estimates.

**Fig. 4:**
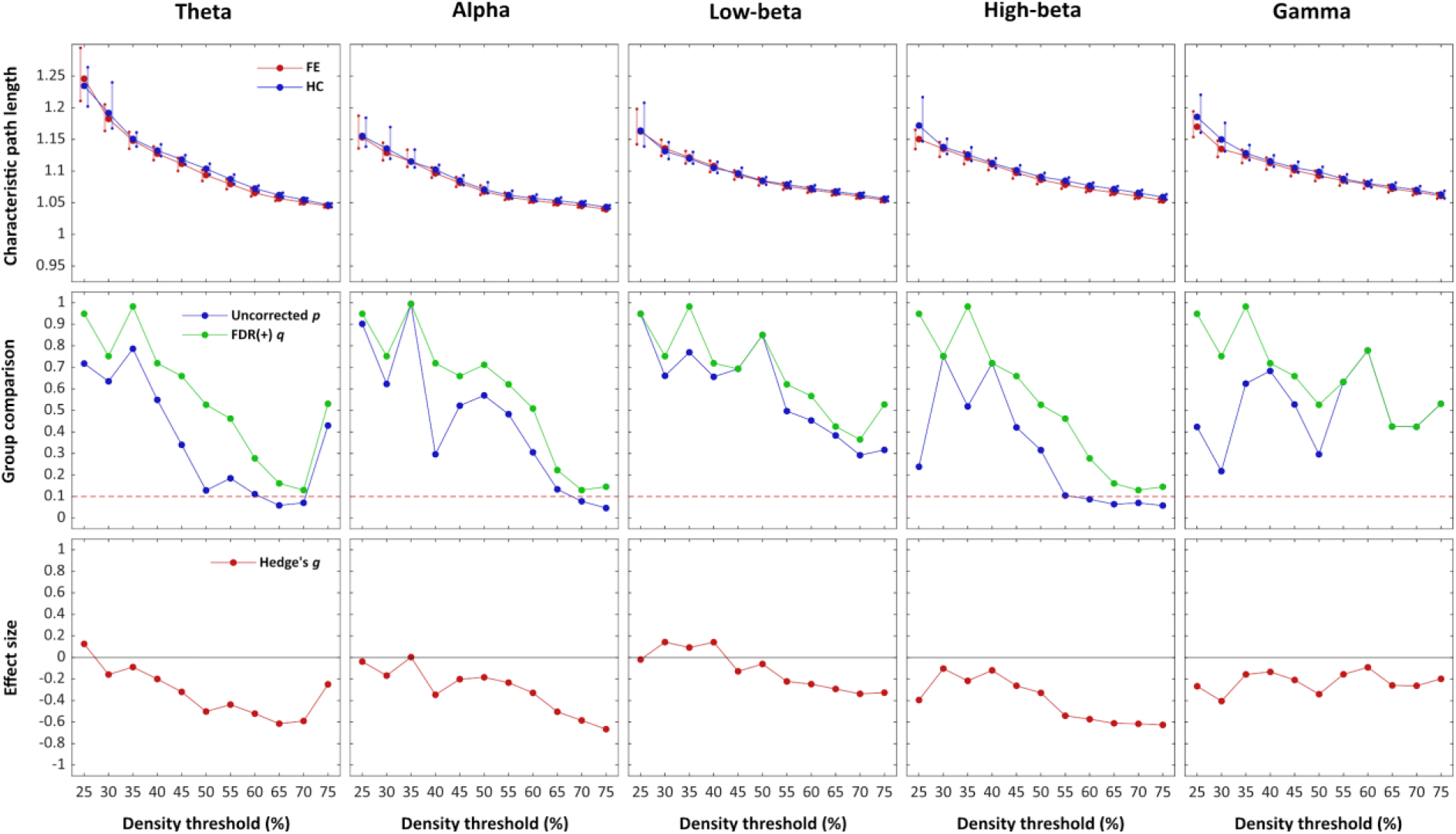
Normalised characteristic path length. Group comparison of the network-wide normalised characteristic path length across DT levels. *Top row:* Normalised characteristic path length, where the vertical lines represent the 95% BCa confidence interval of the mean. *Middle row:* Uncorrected *p* values and FDR adjusted *q* values associated with the permutation tests. The horizontal dashed line represents the critical alpha threshold (*q* < 0.1). *Bottom row:* Effect size estimates.

For all assessed global metrics, the normalised values shifted closer to random with increased network density. Also, the results demonstrated a tendency towards increased effect sizes in the upper end of the DT range.

### 3.2 Local graph metrics

Analyses on the local level revealed stable differences (significant effects in three consecutive DT increments) in several nodes across multiple frequency bands and graph metrics (see Fig. 5). Here, we focus on effects in two nodes: The *left central region* (LCR; node 1 in Fig. 1) and the *left parietal lateral surface* (LPL; node 13). These nodes displayed specific alterations in the alpha band on characteristic path length, node strength and eigenvector centrality (Fig. 6), all three characteristics implied in the evaluation of *hubness* (see discussion). The effects reported in the following are considered statistically significant, having survived FDR adjustment. Table 3 contains a list of all across-DT stable nodal group differences.

**Table 3:**
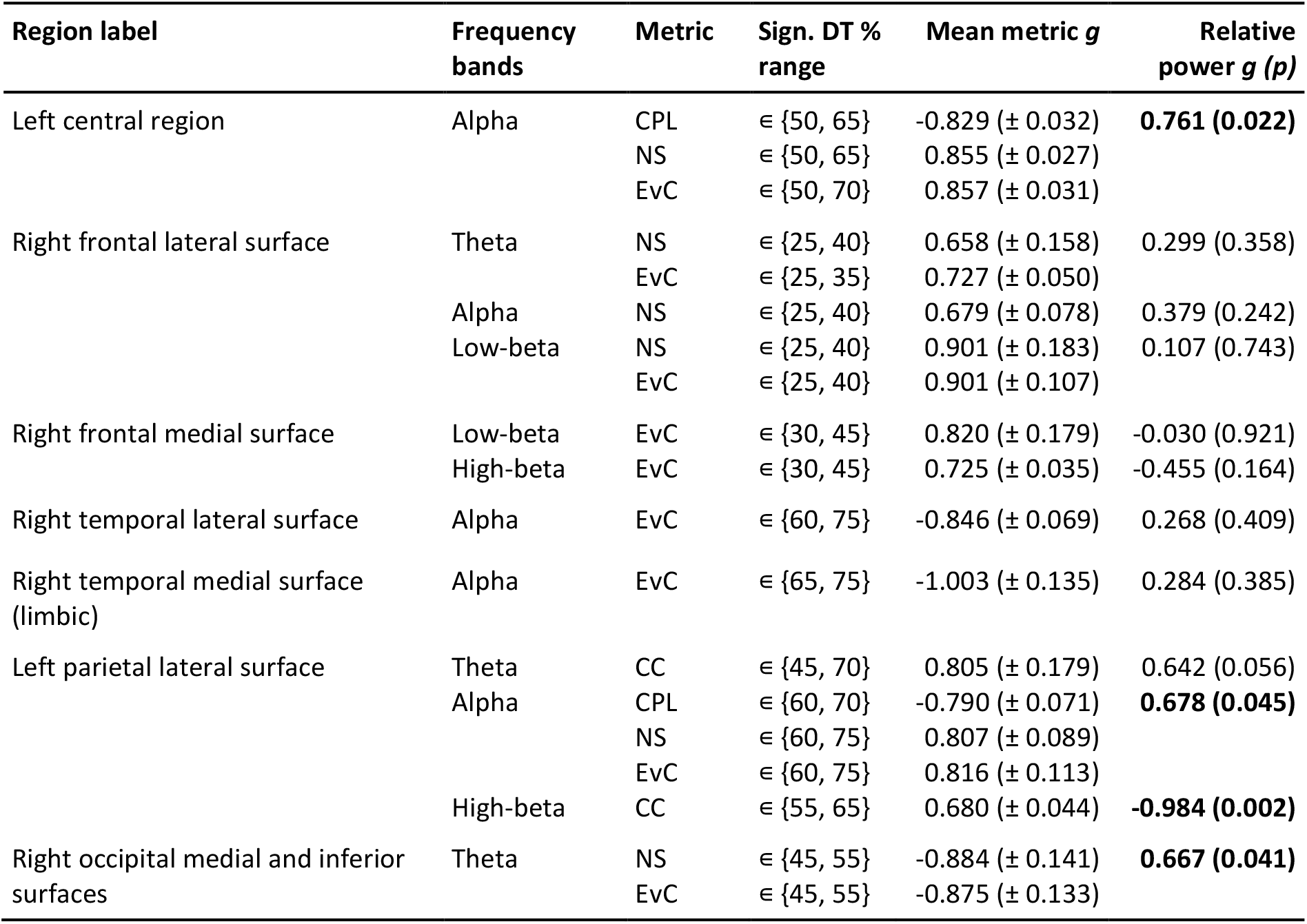
Stable (across three consecutive DT levels) group differences in local graph metrics. The metrics are characteristic path length (CPL), clustering coefficient (CC), node strength (NS) and eigenvector centrality (EvC). The values in the column “Mean metric *g*” are calculated across the significant DT levels. The “Relative power *g (p)*” column provides the frequency band/region group comparison statistics (uncorrected *p* values; significant differences at *p* < 0.05 level in bold).

| Region label | Frequency bands | Metric | Sign. DT % range | Mean metric $g$ | Relative power $g(p)$ |
| --- | --- | --- | --- | --- | --- |
| Left central region | Alpha | CPL | ∈ {50, 65} | -0.829 (± 0.032) | **0.761 (0.022)** |
|  |  | NS | ∈ {50, 65} | 0.855 (± 0.027) |  |
|  |  | EvC | ∈ {50, 70} | 0.857 (± 0.031) |  |
| Right frontal lateral surface | Theta | NS | ∈ {25, 40} | 0.658 (± 0.158) | 0.299 (0.358) |
|  |  | EvC | ∈ {25, 35} | 0.727 (± 0.050) |  |
|  | Alpha | NS | ∈ {25, 40} | 0.679 (± 0.078) | 0.379 (0.242) |
|  | Low-beta | NS | ∈ {25, 40} | 0.901 (± 0.183) | 0.107 (0.743) |
|  |  | EvC | ∈ {25, 40} | 0.901 (± 0.107) |  |
| Right frontal medial surface | Low-beta | EvC | ∈ {30, 45} | 0.820 (± 0.179) | -0.030 (0.921) |
|  | High-beta | EvC | ∈ {30, 45} | 0.725 (± 0.035) | -0.455 (0.164) |
| Right temporal lateral surface | Alpha | EvC | ∈ {60, 75} | -0.846 (± 0.069) | 0.268 (0.409) |
| Right temporal medial surface (limbic) | Alpha | EvC | ∈ {65, 75} | -1.003 (± 0.135) | 0.284 (0.385) |
| Left parietal lateral surface | Theta | CC | ∈ {45, 70} | 0.805 (± 0.179) | 0.642 (0.056) |
|  | Alpha | CPL | ∈ {60, 70} | -0.790 (± 0.071) | **0.678 (0.045)** |
|  |  | NS | ∈ {60, 75} | 0.807 (± 0.089) |  |
|  |  | EvC | ∈ {60, 75} | 0.816 (± 0.113) |  |
|  | High-beta | CC | ∈ {55, 65} | 0.680 (± 0.044) | **-0.984 (0.002)** |
| Right occipital medial and inferior surfaces | Theta | NS | ∈ {45, 55} | -0.884 (± 0.141) | **0.667 (0.041)** |
|  |  | EvC | ∈ {45, 55} | -0.875 (± 0.133) |  |

**Fig. 5:**
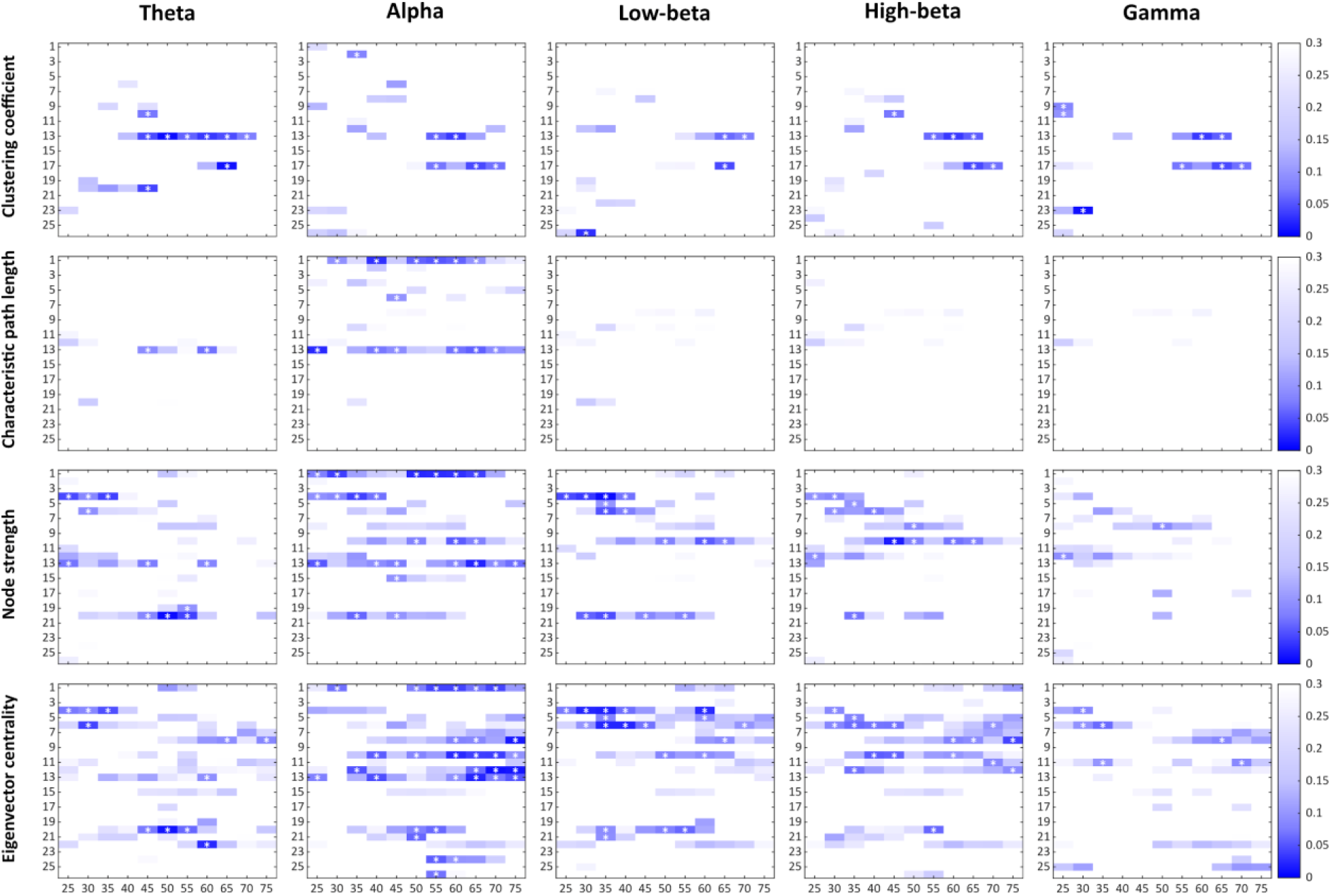
Mass testing of local graph metrics. FDR *q* values from the mass permutation testing of group differences in local graph metrics. Significant (*q* < 0.1) difference in a node is flagged with a white asterisk.

**Fig. 6:**
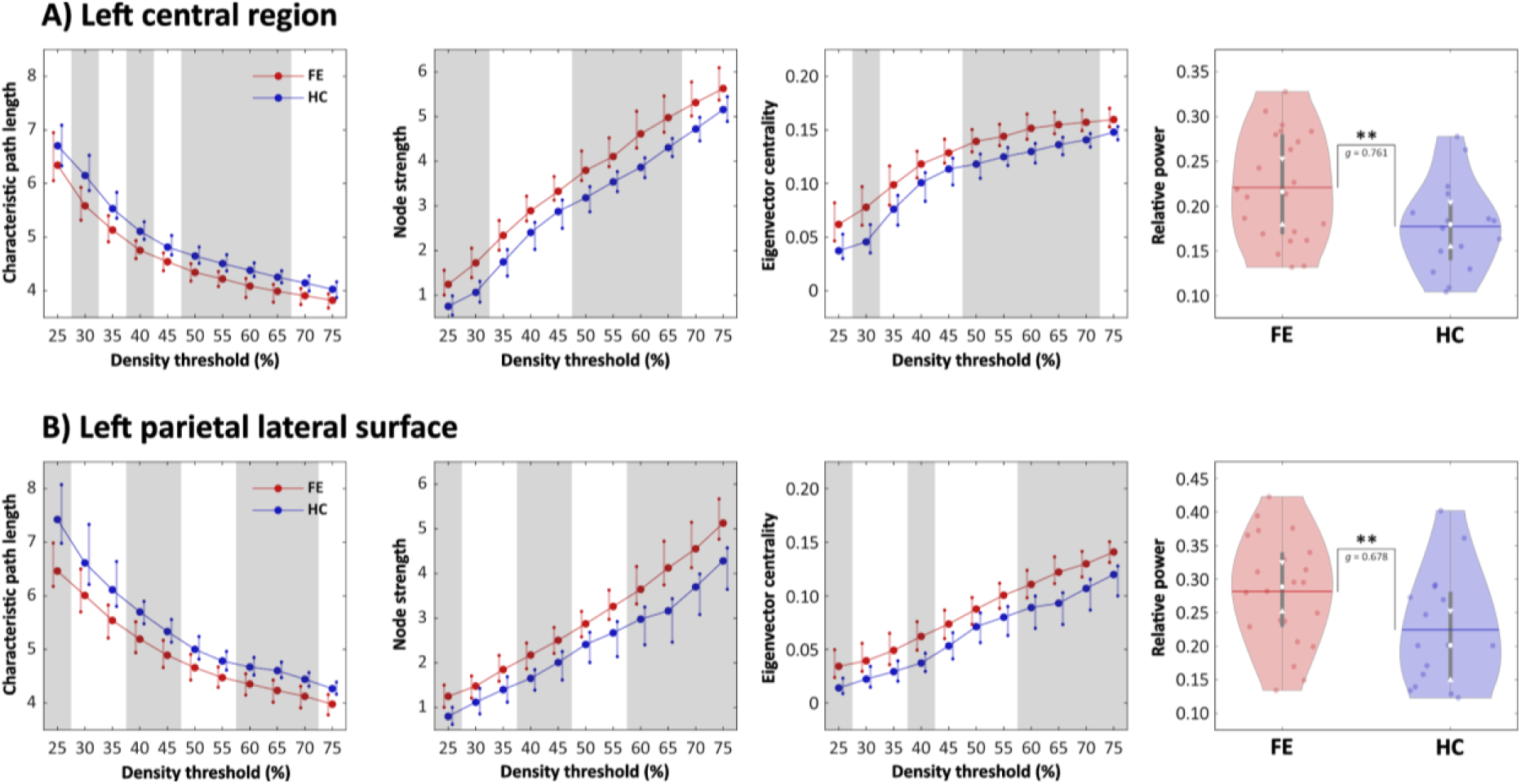
Node hubness. Comparison of local graph metrics and relative power in the LCR (A) and LPL (B) nodes. *Columns 1-3:* Shaded grey areas indicate significant difference at given DT level (*q* < 0.10). *Column 4:* Double asterisk indicates significant group difference when both uncorrected (*p* < 0.05) and FDR adjusted (*q* < 0.10).

For the patient group, the characteristic path length in the alpha band showed stable decrease in both the LCR (DT ∈ {50, 65}, *q* ∈ {0.052, 0.098}, *g* ∈ {−0.858, −0.786}) and LPL (DT ∈ {60, 70}, *q* ∈ {0.058, 0.090}, *g* ∈ {−0.871, −0.737}). Furthermore, in the same band, node strength (LCR: DT ∈ {50, 65}, *q* ∈ {0.025, 0.039}, *g* ∈ {0.820, 0.884}; LPL: DT ∈ {60, 75}, *q* ∈ {0.023, 0.090}, *g* ∈ {0.708, 0.921}) and eigenvector centrality (LCR: DT ∈ {50, 70}, *q* ∈ {0.041, 0.077}, *g* ∈ {0.826, 0.893}; LPL: DT ∈ {60, 75}, *q* ∈ {0.024, 0.094}, *g* ∈ {0.689, 0.962}) were both increased in these nodes in the patient group.

In addition, in the LPL node, increased clustering coefficient was observed in the patient group in the theta (DT ∈ {45, 70}, *q* ∈ {0.009, 0.077}, *g* ∈ {0.596, 1.087}) and high-beta (DT ∈ {55, 65}, *q* ∈ {0.035, 0.063}, *g* ∈ {0.649, 0.730}) bands.

### 3.3 Spectral power

In a *post hoc* approach, we analysed potential group differences in region-wise relative power at the nodes which revealed stable group effects in local graph metrics (Fig. 6, rightmost column). This was done on the basis of delineating potential confounding effects of power on graph metrics (see discussion on source leakage). The below reported *p* values are uncorrected with regard to multiple comparisons.

For the patient group, increased alpha power was observed in both LCR (*p* = 0.022, *g* = 0.761) and LPL (*p* = 0.045, *g* = 0.678) nodes. The LPL node also showed a significant decrease for the patient group in high-beta power (*p* = 0.002, *g* = −0.984). In addition, in the node representing the *right occipital and medial inferior surfaces*, the patient group displayed an increase in relative power (*p* = 0.041, *g* = 0.667).

The full analysis of all regions across all frequency bands is presented in Fig. 7.

**Fig. 7:**
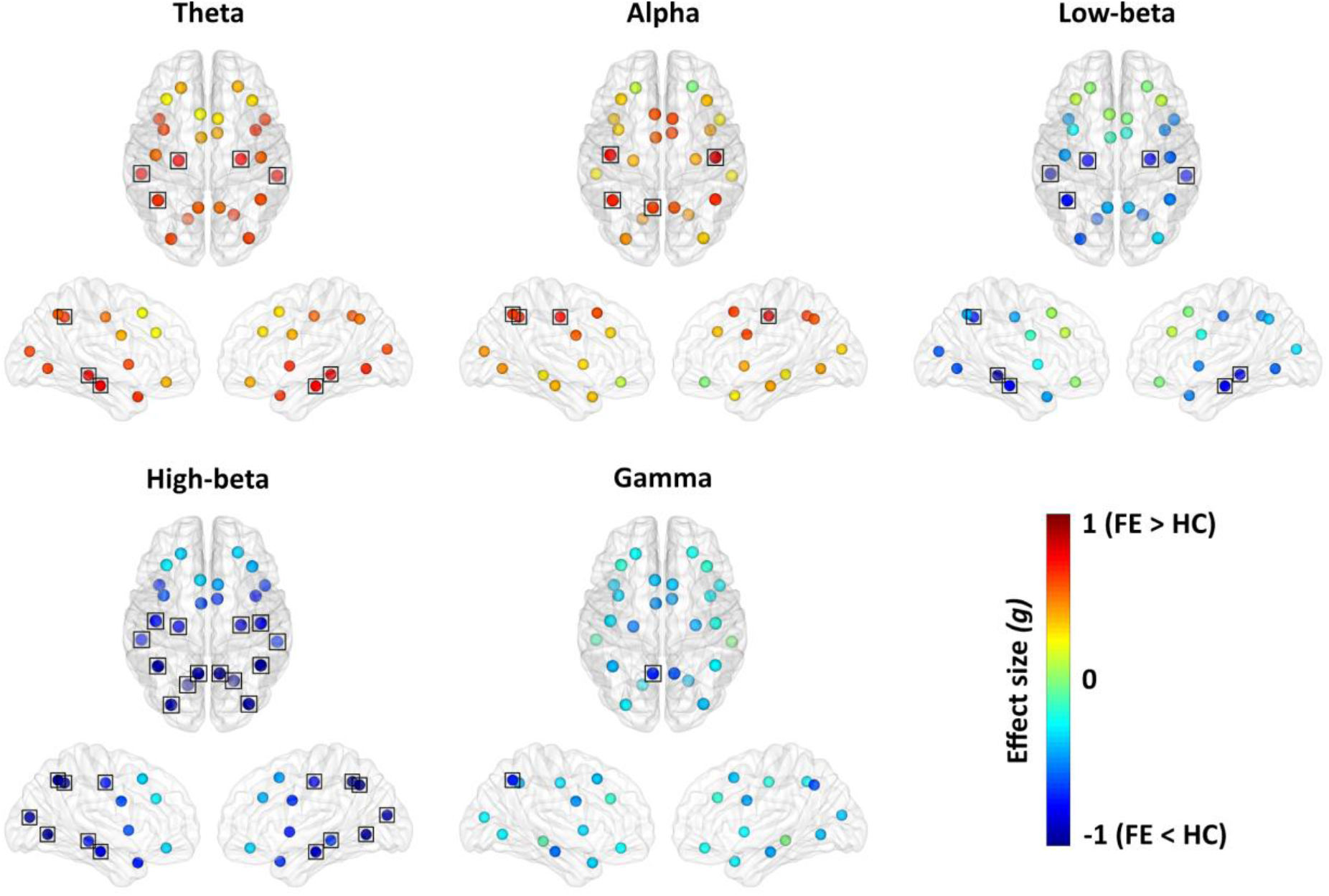
Spectral power. Region-wise group comparison of relative power. A black rectangle indicates significance at *q* < 0.10 level after FDR adjustment of *p* values from the permutation test.

## 4 Discussion

To our knowledge, the current study is the first to present a comprehensive analysis of both global and local graph metrics in adult focal epilepsy via FC estimated from source-space scalp EEG. In recent years, analysis of functional brain networks has shown clinical potential in the diagnostics and treatment of epilepsy. Here, we demonstrate that changes to functional network topology on both the global and local levels are evident even in well-functioning patients, suggesting that altered network organisation is a stable trait of focal epilepsy. In our results, epilepsy patients showed consistently higher small world indices, and the investigation of individual nodes revealed evidence of altered hubness of the left central region and the left parietal lateral surface. In the following, we will discuss the clinical relevance of our findings to the understanding of focal epilepsy.

### 4.1 Global metrics: Increased small worldness in focal epilepsy

Network-wide changes in epilepsy are consistently reported, yet their clinical significance remains inadequately understood. It is generally accepted that epileptic symptoms arise from abnormal neuronal synchronisation and excitability (Engel et al. 2013). Although these phenomena can be observed experimentally through FC, the understanding of their underlying mechanisms remains scarce (van Diessen et al. 2013). Graph theoretical characteristics of topological network organisation could remedy this shortcoming (Bernhardt et al. 2015). However, the links between graph metrics and the pathophysiological mechanisms of epilepsy have yet to be firmly established.

Here, we observed an increase in the small world index of the theta, alpha, high-beta and gamma bands in the patients relative to controls. These observations partly corroborate previous reports, where focal epilepsy patients have displayed increased small world index in the theta and alpha bands (Quraan et al. 2013). Interestingly, the shift towards increased small worldness contrasts earlier findings suggesting that epileptic activity is associated with increased regularisation of brain networks, manifested as a dual increase in clustering coefficient and characteristic path length (Horstmann et al. 2010; Vecchio et al. 2015). The latter tendency was also reported in an across-modality meta-analysis (van Diessen et al. 2014), and in an fMRI study by Pedersen and colleagues (2015), who suggested that epileptic network activity may arise in accordance with the *fault tolerant network* (Johnson 1984) hypothesis. In this scenario, a network with the potential of evolving dysfunctional nodes will compensate by altering its topology towards increased regularity. This shift in favour of network regularisation, however, was not evident in our results, which in contrast demonstrated a decrease in characteristic path length. The reason for this discrepancy is not clear; however, both the referenced EEG studies differed methodologically from ours, in where one (Vecchio et al. 2015) focused exclusively on fronto-temporal areas in dense weighted network matrices, whereas the other (Horstmann et al. 2010) employed sensor-space connectivity estimates, thus more affected by volume conduction confounds. Notably, another study examined the temporal lobe specifically with intracranial EEG, and found that small worldness in focal epilepsy could depend on the time since initial diagnosis (van Dellen et al. 2009), an association that was not examined here.

However, the relevance of the small world index remains an issue of debate (Papo et al. 2016). As a summary metric, it captures the network topology’s trade-off between local segregation and global integration, *i*.*e*., to what degree the brain retains its modular functional organisation with a high number of short communication paths, yet facilitates signal transmission between modules via long-range paths to achieve integrated functioning (Bertolero et al. 2015). Mathematically, the index can increase either as a function of an increased clustering coefficient, a decreased characteristic path length, or both; however, it is unclear in what range the brain can reside and still retain its optimal functioning. From our data, an association between hypersynchronous neuronal activity and increased small worldness in epilepsy is likely. Hypersynchronous neuronal activity is classically understood as a “problem of regions and neurons connected or communicating too readily” (Kramer and Cash 2012). More importantly, in focal epilepsies without secondary generalisation, such hypersynchronous activity originates in a delimited brain region with propagation restricted to other regions with high (functional) proximity to the epileptogenic zone. Thus, high FC between neighbouring regions will be likely to occur, and the proportional threshold scheme will result in the rejection of more long-range connections in the patient group relative to the controls. This notion is supported further by evidence that global efficiency (inverse characteristic path length) is higher in patients with generalised epilepsy than focal epilepsy, which again is higher than in healthy controls (Niso et al. 2015).

### 4.2 Local metrics: Altered network hubs

Node-level graph metrics are considerably less discussed in the literature than global metrics, and the interpretational framework for local network alterations remains limited. However, the concept of *hubness* (*i*.*e*., the degree to which a node constitutes a hub in the network) has gained some influence (van den Heuvel et al. 2010; Ridley et al. 2015; Lee et al. 2018). A *hub* is a node of relatively high importance to the network, being essential to efficient integration of information across the network. Consequently, local disruptions of hub nodes are more likely to cause severe impairment to global integrative processes and network organisation, due to their central positions (van den Heuvel and Sporns 2013), than disruptions to regular nodes. In terms of graph metrics, a hub node may be characterised by high node degree/strength, high centrality, and short characteristic path length, combined with low clustering coefficient (van den Heuvel et al. 2010).

The role of (changing) brain network hubs has yet to be discerned in the context of focal epilepsy. Provided that brain hubs are regions with a strong degree of participation in information transmission throughout the network, they must necessarily also be considered at risk to act as bottlenecks in the system (Marois and Ivanoff 2005; van den Heuvel and Sporns 2013). Pursuing this notion, increased hubness in a selection of nodes in epilepsy might imply that epileptic activity reduces the efficiency of brain networks by excessively routing information through these nodes, maladaptively generating communication bottlenecks. Importantly, in this scenario, a dysfunction in information flow would be an effect of epileptic neuronal activity, and a possible mediator for cognitive dysfunction and other non-seizure symptoms of epilepsy. In theory, such nodes could serve as potential targets for surgical intervention; however, their anatomical localisation might render this utility impractical.

Here, we demonstrate a shift towards increased hubness in the left central region and the left parietal lateral surface in the alpha band of focal epilepsy patients. This shift manifests as nodal changes characterised by shorter characteristic path length and increased node strength and eigenvector centrality. Interestingly, the parietal lateral surface, as defined here, includes the angular gyrus, a region in which local network alterations have been reported previously (Ridley et al. 2015). Furthermore, the angular gyrus is recognised as (part of) a functional hub of the default mode network (DMN), implicated in introspective and social cognitive processes (Andrews-Hanna et al. 2014), and dysfunctions of the DMN have been demonstrated interictally in focal epilepsy (Burianová et al. 2017; Lee et al. 2018; Ofer et al. 2019). To our knowledge, the current study is the first to do so using source-space analysis based on EEG, thus advancing the recognition of specific dysfunctional networks and brain regions. Moreover, temporal lobe epilepsies are associated with dysfunctional theory of mind and facial emotion recognition (Bora and Meletti 2016), which are social cognition processes dependent on the DMN.

### 4.3 Across-threshold stability

A subsidiary aim of the current effort was to probe the stability of group differences in graph metrics across a predefined range of adjacency matrix densities, a matter which remains unresolved in the field. Functional networks are per definition arbitrary, and no basis exists on which to define an ecologically valid threshold level. The motivation to impose a threshold on the FC matrix prior to computing graph metrics, comes from the assumption that weak connections are more likely to be spurious (and thus conceal key topological properties) than strong connections (Fornito et al. 2012; for a discussion of PLV, see also Aydore et al. 2013).

Mathematically, network density directly influences a range of graph metrics (for a detailed account, see van Wijk et al. 2010). The primary concerns relating to this issue are the reproducibility of findings across studies and the comparison of graph metrics across groups within the same study. The latter issue is mostly solved by the application of *proportional* (*i*.*e*., density-based) thresholds, where the connection weight cut-off varies individually in order to standardise the network density across subjects. Here, we corroborate previous reports that the proportional threshold scheme produces relatively consistent group differences across the threshold continuum (Quraan et al. 2013; van den Heuvel et al. 2017). However, in contrast to absolute thresholds, the use of proportional thresholds does not exclude the possibility that differences in overall FC of the dense matrices influences the resulting graph metrics (van den Heuvel et al. 2017). Our results did not reveal altered overall FC between groups, suggesting that the topological group effects were not inflated by the network threshold scheme.

We investigated group differences across a middle range of density threshold levels, thus excluding the extremes, which have been shown to produce erroneous results in fMRI-based connectivity (Garrison et al. 2015). In accordance with previous reports, we found relatively stable directionality of effects across density levels (*i*.*e*., there were few “sign reversals”), when a proportional threshold scheme was used (Garrison et al. 2015; van den Heuvel et al. 2017). It should be noted that in the current approach, in contrast to the referenced studies, network matrices were constrained by the requirement to remain fully connected after threshold imposition.

### 4.4 Clinical relevance of network analysis in epilepsy and future aims

Epilepsy is increasingly considered a disease of brain network pathology, and procedures utilising FC and graph theory have been proposed for several clinical applications relating to epilepsy diagnostics and treatment (Stefan and Lopes da Silva 2013; van Mierlo et al. 2019). Among the most investigated are potential improvements to pre-surgical evaluation, such as localisation of epileptogenic zones (Panzica et al. 2013; van Mierlo et al. 2014), and prediction of resection outcome. An intriguing example of the latter, is a recently introduced method of virtual resection of functional brain networks derived from intracranial EEG (Kini et al. 2019). The authors report high precision in predicting which patients with drug-resistant focal epilepsy will benefit from epilepsy surgery, as well as in restricting the resection zone. In terms of non-invasive methods to improve epilepsy surgery, FC from source EEG shows promise in identification of surgical targets (Staljanssens et al. 2017), and graph metrics (in this case derived from fMRI) have been successfully employed to predict the cognitive outcome for patients undergoing temporal lobe resections (Doucet et al. 2015). The association between cognitive functioning and network metrics has also been demonstrated with EEG methods, in cohorts of Alzheimer’s disease and mild cognitive impairment patients (Vecchio et al. 2016), and in healthy subjects (Langer et al. 2012). Here, we observed robust network alterations in theta, alpha, high-beta and gamma bands; all oscillation frequencies associated with different cognitive functions. Both slow-wave delta and theta, and gamma frequencies have been consistently implicated in memory processes (Raghavachari et al. 2006; Axmacher et al. 2008; Sauseng et al. 2009; Jacobs and Kahana 2010; Hanslmayr et al. 2019), whereas alpha rhythm is associated with control of attention (Mathewson et al. 2009); both of which are affected in focal epilepsy (Hermann et al. 2007). However, whether aberrant network topology in certain oscillatory bands is related to dysfunction in specific cognitive domains, remains unclear, and should constitute an important target for future studies.

In sum, it is reasonable to believe that EEG-based functional network analysis in the near future will play an integral part in diagnostics and treatment of epilepsy. As demonstrated here, although the patient cohort was characterised by successful ASM therapy and high quality of life, functional network alterations were still present relative to non-epileptic peers. Thus, an aim for future studies should be to investigate specific topographical alterations in relation to customary elements of epilepsy diagnostics and treatment, such as cognitive dysfunction and ASM. Provided the small sample investigated in the current study, we did not stratify the patients including information about medication. However, some efforts have been made towards examining the potential of network analysis as an indicator for successful treatment, although clinical validation remains sparse. One study found that carbamazepine and oxcarbazepine affected the betweenness centrality of several hubs, suggesting that these medications work, at least in part, by altering (or reverting the alterations introduced by the disease) the brain’s hub organisation (Haneef et al. 2015). Another study found selective differences in clustering coefficient and global efficiency in patients using topiramate compared to other medications (van Veenendaal et al. 2017). Taken together, these studies suggest that ASMs may have differential effects on functional brain networks. If this holds true, network topology might guide the choice of ASM administered to newly diagnosed epilepsies, an important goal for clinical neurology. Thus, comprehensive studies of ASMs’ effect on functional network topology are highly warranted.

### 4.5 Limitations

Some limitations with the current study should be addressed. First, our sample sizes were small. Although we were able to identify significant group differences, the low number of participants did not allow precise stratification with regard to individual clinical factors, such as ASM therapy, epileptogenic zone localisation and structural brain lesions. The current sample size also limits the generalisability of our statistical approach; thus, we encourage future studies to corroborate the current findings in larger samples Second, compared to fMRI and MEG, EEG has limitations regarding spatial resolution. We have mitigated this shortcoming by employing high-quality source reconstruction, and by using a reduced version of the AAL atlas with larger regions. On the other hand, the low cost and clinical availability associated with EEG justify this trade-off.

Third, electrophysiologically derived FC is limited by the effect of source leakage. If the leakage varies across groups, differences can arise in the FC matrices, and by extension, the graph metrics. While some FC measures eliminate the effects of leakage by removing zero-lag synchronisation (Ewald et al. 2012; Bruña et al. 2018), these show low test-retest reliability (Colclough et al. 2016; Garcés et al. 2016). Here, we decided to quantify source leakage, using local power as a proxy (Muthukumaraswamy and Singh 2011). In our data, local power in the alpha band was increased in the patients in both LCR and LPL nodes. This could result from higher leakage, increasing the FC with neighbouring areas; however, the associated effect sizes were smaller than those of the graph metrics, arguing that the differences in power were less relevant. In further support of this notion, the remaining nodes displaying stable alterations did not show significant differences in power. Overall, the power differences between groups are most pronounced in the high-beta band (Fig. 6), in which we did not observe changes in node hubness.

### 4.6 Conclusion

Although we investigated a relatively small sample, we have presented evidence that focal epilepsy patients, despite good seizure control and high quality of life, exhibit widespread functional network alterations relative to healthy peers. Interestingly, these discrepancies were evident at the group-level, suggesting that functional network alterations are evident in focal epilepsy regardless of (or as the sum of) individual clinical factors. Importantly, such findings highlight the clinical application of network analysis as a potential framework in which secondary epilepsy symptoms, such as cognitive dysfunction, and ASM therapy may be understood.

## Data Availability

The data that support the findings of this study are available from the corresponding author (or his affiliated institution), upon reasonable request.

## Abbreviations

AAL: Automated Anatomical Labeling
ASM: anti-seizure medication
DMN: default mode network
DT: density threshold
FC: functional connectivity
FDR: false discovery rate
FE: focal epilepsy
HC: healthy control
LCR: left central region
LPL: left parietal lateral
MST: minimum spanning tree
PLV: phase-locking value

## References

Andrews-Hanna JR, Smallwood J, Spreng RN. The default network and self-generated thought: component processes, dynamic control, and clinical relevance. Ann N Y Acad Sci [Internet]. 2014 May;1316:29–52. Available from: http://dx.doi.org/10.1111/nyas.12360

Ashburner J, Friston KJ. Unified segmentation. Neuroimage [Internet]. 2005 Jul 1;26(3):839–51. Available from: http://dx.doi.org/10.1016/j.neuroimage.2005.02.018

Axmacher N, Elger CE, Fell J. Ripples in the medial temporal lobe are relevant for human memory consolidation. Brain [Internet]. 2008 Jul;131(Pt 7):1806–17. Available from: http://dx.doi.org/10.1093/brain/awn103

Aydore S, Pantazis D, Leahy RM. A note on the phase locking value and its properties. Neuroimage [Internet]. 2013 Jul 1;74:231–44. Available from: http://dx.doi.org/10.1016/j.neuroimage.2013.02.008

Bastos AM, Schoffelen J-M. A Tutorial Review of Functional Connectivity Analysis Methods and Their Interpretational Pitfalls. Front Syst Neurosci [Internet]. 2015;9:175. Available from: http://dx.doi.org/10.3389/fnsys.2015.00175

Behrens TEJ, Sporns O. Human connectomics. Curr Opin Neurobiol [Internet]. 2012 Feb;22(1):144–53. Available from: http://dx.doi.org/10.1016/j.conb.2011.08.005

Belouchrani A, Abed-Meraim K, Cardoso JF, Moulines E. Second-order blind separation of temporally correlated sources. In: Proc Int Conf Digital Signal Processing [Internet]. Citeseer; 1993. p. 346–51. Available from: https://www.researchgate.net/profile/Adel_Belouchrani/publication/2699542_Second_Order_Blind_Separation_of_Temporally_Correlated_Sources/links/00463517ab3e0aed06000000/Second-Order-Blind-Separation-of-Temporally-Correlated-Sources.pdf

Benjamini Y, Yekutieli D. The Control of the False Discovery Rate in Multiple Testing under Dependency. Ann Stat [Internet]. 2001;29(4):1165–88. Available from: http://www.jstor.org/stable/2674075

Bernhardt BC, Bonilha L, Gross DW. Network analysis for a network disorder: The emerging role of graph theory in the study of epilepsy. Epilepsy Behav [Internet]. 2015 Sep;50:162–70. Available from: http://dx.doi.org/10.1016/j.yebeh.2015.06.005

Bertolero MA, Yeo BTT, D’Esposito M. The modular and integrative functional architecture of the human brain. Proc Natl Acad Sci U S A [Internet]. 2015 Dec 8;112(49):E6798–807. Available from: http://dx.doi.org/10.1073/pnas.1510619112

Besserve M, Martinerie J, Garnero L. Improving quantification of functional networks with EEG inverse problem: evidence from a decoding point of view. Neuroimage [Internet]. 2011 Apr 15;55(4):1536–47. Available from: http://dx.doi.org/10.1016/j.neuroimage.2011.01.056

Bigdely-Shamlo N, Mullen T, Kothe C, Su K-M, Robbins KA. The PREP pipeline: standardized preprocessing for large-scale EEG analysis. Front Neuroinform [Internet]. 2015 Jun 18;9(16):16. Available from: http://dx.doi.org/10.3389/fninf.2015.00016

Bora E, Meletti S. Social cognition in temporal lobe epilepsy: A systematic review and meta-analysis. Epilepsy Behav [Internet]. 2016 Jul;60:50–7. Available from: http://dx.doi.org/10.1016/j.yebeh.2016.04.024

Bruña R, Maestú F, Pereda E. Phase locking value revisited: teaching new tricks to an old dog. J Neural Eng [Internet]. 2018 Oct;15(5):056011. Available from: http://dx.doi.org/10.1088/1741-2552/aacfe4

Brunner C, Billinger M, Seeber M, Mullen TR, Makeig S. Volume Conduction Influences Scalp-Based Connectivity Estimates. Front Comput Neurosci [Internet]. 2016 Nov 22;10:121. Available from: http://dx.doi.org/10.3389/fncom.2016.00121

Burianová H, Faizo NL, Gray M, Hocking J, Galloway G, Reutens D. Altered functional connectivity in mesial temporal lobe epilepsy. Epilepsy Res [Internet]. 2017 Nov;137:45–52. Available from: http://dx.doi.org/10.1016/j.eplepsyres.2017.09.001

de Cheveigné A. ZapLine: A simple and effective method to remove power line artifacts. Neuroimage [Internet]. 2020 Feb 15;207:116356. Available from: http://dx.doi.org/10.1016/j.neuroimage.2019.116356

Colclough GL, Woolrich MW, Tewarie PK, Brookes MJ, Quinn AJ, Smith SM. How reliable are MEG resting-state connectivity metrics? Neuroimage [Internet]. 2016 Sep;138:284–93. Available from: http://dx.doi.org/10.1016/j.neuroimage.2016.05.070

van Dellen E, Douw L, Baayen JC, Heimans JJ, Ponten SC, Vandertop WP, et al. Long-term effects of temporal lobe epilepsy on local neural networks: a graph theoretical analysis of corticography recordings. PLoS One [Internet]. 2009 Nov 26;4(11):e8081. Available from: http://dx.doi.org/10.1371/journal.pone.0008081

Delorme A, Makeig S. EEGLAB: an open source toolbox for analysis of single-trial EEG dynamics including independent component analysis. J Neurosci Methods [Internet]. 2004 Mar 15;134(1):9–21. Available from: http://dx.doi.org/10.1016/j.jneumeth.2003.10.009

van Diessen E, Diederen SJH, Braun KPJ, Jansen FE, Stam CJ. Functional and structural brain networks in epilepsy: what have we learned? Epilepsia [Internet]. 2013 Nov;54(11):1855–65. Available from: http://dx.doi.org/10.1111/epi.12350

van Diessen E, Numan T, van Dellen E, van der Kooi AW, Boersma M, Hofman D, et al. Opportunities and methodological challenges in EEG and MEG resting state functional brain network research. Clin Neurophysiol [Internet]. 2015 Aug;126(8):1468–81. Available from: http://dx.doi.org/10.1016/j.clinph.2014.11.018

van Diessen E, Zweiphenning WJEM, Jansen FE, Stam CJ, Braun KPJ, Otte WM. Brain Network Organization in Focal Epilepsy: A Systematic Review and Meta-Analysis. PLoS One [Internet]. 2014 Dec 10;9(12):e114606. Available from: http://dx.doi.org/10.1371/journal.pone.0114606

Doucet GE, Rider R, Taylor N, Skidmore C, Sharan A, Sperling M, et al. Presurgery resting-state local graph-theory measures predict neurocognitive outcomes after brain surgery in temporal lobe epilepsy. Epilepsia [Internet]. 2015 Apr;56(4):517–26. Available from: http://dx.doi.org/10.1111/epi.12936

Douw L, van Dellen E, Gouw AA, Griffa A, de Haan W, van den Heuvel M, et al. The road ahead in clinical network neuroscience. Netw Neurosci [Internet]. 2019 Sep 1;3(4):969–93. Available from: http://dx.doi.org/10.1162/netn_a_00103

Elshahabi A, Klamer S, Sahib AK, Lerche H, Braun C, Focke NK. Magnetoencephalography Reveals a Widespread Increase in Network Connectivity in Idiopathic/Genetic Generalized Epilepsy. PLoS One [Internet]. 2015 Sep 14;10(9):e0138119. Available from: http://dx.doi.org/10.1371/journal.pone.0138119

Engel J Jr, Thompson PM, Stern JM, Staba RJ, Bragin A, Mody I. Connectomics and epilepsy. Curr Opin Neurol [Internet]. 2013 Apr;26(2):186–94. Available from: http://dx.doi.org/10.1097/WCO.0b013e32835ee5b8

Engels MMA, Stam CJ, van der Flier WM, Scheltens P, de Waal H, van Straaten ECW. Declining functional connectivity and changing hub locations in Alzheimer’s disease: an EEG study. BMC Neurol [Internet]. 2015 Aug 20;15(145):145. Available from: http://dx.doi.org/10.1186/s12883-015-0400-7

Ewald A, Marzetti L, Zappasodi F, Meinecke FC, Nolte G. Estimating true brain connectivity from EEG/MEG data invariant to linear and static transformations in sensor space. Neuroimage [Internet]. 2012 Mar;60(1):476–88. Available from: http://dx.doi.org/10.1016/j.neuroimage.2011.11.084

Fan J, Xu P, Van Dam NT, Eilam-Stock T, Gu X, Luo Y-J, et al. Spontaneous brain activity relates to autonomic arousal. J Neurosci [Internet]. 2012 Aug 15;32(33):11176–86. Available from: http://dx.doi.org/10.1523/JNEUROSCI.1172-12.2012

Fornito A, Zalesky A, Pantelis C, Bullmore ET. Schizophrenia, neuroimaging and connectomics. Neuroimage [Internet]. 2012 Oct 1;62(4):2296–314. Available from: http://dx.doi.org/10.1016/j.neuroimage.2011.12.090

Garcés P, Martín-Buro MC, Maestú F. Quantifying the Test-Retest Reliability of Magnetoencephalography Resting-State Functional Connectivity. Brain Connect [Internet]. 2016 Jul;6(6):448–60. Available from: http://dx.doi.org/10.1089/brain.2015.0416

Garrison KA, Scheinost D, Finn ES, Shen X, Constable RT. The (in)stability of functional brain network measures across thresholds. Neuroimage [Internet]. 2015 Sep;118:651–61. Available from: http://dx.doi.org/10.1016/j.neuroimage.2015.05.046

Genovese CR, Lazar NA, Nichols T. Thresholding of statistical maps in functional neuroimaging using the false discovery rate. Neuroimage [Internet]. 2002 Apr;15(4):870–8. Available from: http://dx.doi.org/10.1006/nimg.2001.1037

Gramfort A, Papadopoulo T, Olivi E, Clerc M. OpenMEEG: opensource software for quasistatic bioelectromagnetics. Biomed Eng Online [Internet]. 2010 Sep 6;9:45. Available from: http://dx.doi.org/10.1186/1475-925X-9-45

Hallett M, de Haan W, Deco G, Dengler R, Di Iorio R, Gallea C, et al. Human brain connectivity: Clinical applications for clinical neurophysiology. Clin Neurophysiol [Internet]. 2020 Jul;131(7):1621–51. Available from: http://dx.doi.org/10.1016/j.clinph.2020.03.031

Haneef Z, Chiang S. Clinical correlates of graph theory findings in temporal lobe epilepsy. Seizure [Internet]. 2014 Nov;23(10):809–18. Available from: http://dx.doi.org/10.1016/j.seizure.2014.07.004

Haneef Z, Levin HS, Chiang S. Brain Graph Topology Changes Associated with Anti-Epileptic Drug Use. Brain Connect [Internet]. 2015 Jun;5(5):284–91. Available from: http://dx.doi.org/10.1089/brain.2014.0304

Hanslmayr S, Axmacher N, Inman CS. Modulating Human Memory via Entrainment of Brain Oscillations. Trends Neurosci [Internet]. 2019 Jul;42(7):485–99. Available from: http://dx.doi.org/10.1016/j.tins.2019.04.004

Hassan M, Wendling F. Electroencephalography Source Connectivity: Aiming for High Resolution of Brain Networks in Time and Space. IEEE Signal Process Mag [Internet]. 2018 May;35(3):81–96. Available from: https://ieeexplore.ieee.org/document/8351910/

Hermann B, Seidenberg M, Lee E-J, Chan F, Rutecki P. Cognitive phenotypes in temporal lobe epilepsy. J Int Neuropsychol Soc [Internet]. 2007 Jan;13(1):12–20. Available from: http://dx.doi.org/10.1017/S135561770707004X

van den Heuvel MP, de Lange SC, Zalesky A, Seguin C, Yeo BTT, Schmidt R. Proportional thresholding in resting-state fMRI functional connectivity networks and consequences for patient-control connectome studies: Issues and recommendations. Neuroimage [Internet]. 2017 May 15;152:437–49. Available from: http://dx.doi.org/10.1016/j.neuroimage.2017.02.005

van den Heuvel MP, Mandl RCW, Stam CJ, Kahn RS, Hulshoff Pol HE. Aberrant frontal and temporal complex network structure in schizophrenia: a graph theoretical analysis. J Neurosci [Internet]. 2010 Nov 24;30(47):15915–26. Available from: http://dx.doi.org/10.1523/JNEUROSCI.2874-10.2010

van den Heuvel MP, Sporns O. Network hubs in the human brain. Trends Cogn Sci [Internet]. 2013 Dec;17(12):683–96. Available from: http://dx.doi.org/10.1016/j.tics.2013.09.012

Holmes GL. Cognitive impairment in epilepsy: the role of network abnormalities. Epileptic Disord [Internet]. 2015 Jun;17(2):101–16. Available from: http://dx.doi.org/10.1684/epd.2015.0739

Horstmann M-T, Bialonski S, Noennig N, Mai H, Prusseit J, Wellmer J, et al. State dependent properties of epileptic brain networks: comparative graph-theoretical analyses of simultaneously recorded EEG and MEG. Clin Neurophysiol [Internet]. 2010 Feb;121(2):172–85. Available from: http://dx.doi.org/10.1016/j.clinph.2009.10.013

Huang Y, Parra LC, Haufe S. The New York Head-A precise standardized volume conductor model for EEG source localization and tES targeting. Neuroimage [Internet]. 2016 Oct 15;140:150–62. Available from: http://dx.doi.org/10.1016/j.neuroimage.2015.12.019

Humphries MD, Gurney K. Network “small-world-ness”: a quantitative method for determining canonical network equivalence. PLoS One [Internet]. 2008 Apr 30;3(4):e0002051. Available from: http://dx.doi.org/10.1371/journal.pone.0002051

Jacobs J, Kahana MJ. Direct brain recordings fuel advances in cognitive electrophysiology. Trends Cogn Sci [Internet]. 2010 Apr;14(4):162–71. Available from: http://dx.doi.org/10.1016/j.tics.2010.01.005

Johnson B. Fault-tolerant microprocessor-based systems. IEEE Micro [Internet]. 1984;(6):6–21. Available from: https://www.computer.org/csdl/mags/mi/1984/06/04071150.pdf

King BR, van Ruitenbeek P, Leunissen I, Cuypers K, Heise K-F, Santos Monteiro T, et al. Age-Related Declines in Motor Performance are Associated With Decreased Segregation of Large-Scale Resting State Brain Networks. Cereb Cortex [Internet]. 2018 Dec 1;28(12):4390–402. Available from: http://dx.doi.org/10.1093/cercor/bhx297

Kini LG, Bernabei JM, Mikhail F, Hadar P, Shah P, Khambhati AN, et al. Virtual resection predicts surgical outcome for drug-resistant epilepsy. Brain [Internet]. 2019 Dec 1;142(12):3892–905. Available from: http://dx.doi.org/10.1093/brain/awz303

Kramer MA, Cash SS. Epilepsy as a disorder of cortical network organization. Neuroscientist [Internet]. 2012 Aug;18(4):360–72. Available from: http://dx.doi.org/10.1177/1073858411422754

Lachaux JP, Rodriguez E, Martinerie J, Varela FJ. Measuring phase synchrony in brain signals. Hum Brain Mapp [Internet]. 1999;8(4):194–208. Available from: 3.0.co;2- c”>http://dx.doi.org/10.1002/(sici)1097-0193(1999)8:4<194::aid-hbm4>3.0.co;2-c

Lakens D. Calculating and reporting effect sizes to facilitate cumulative science: a practical primer for t-tests and ANOVAs. Front Psychol [Internet]. 2013 Nov 26;4:863. Available from: http://dx.doi.org/10.3389/fpsyg.2013.00863

Langer N, Pedroni A, Gianotti LRR, Hänggi J, Knoch D, Jäncke L. Functional brain network efficiency predicts intelligence. Hum Brain Mapp [Internet]. 2012 Jun;33(6):1393–406. Available from: http://dx.doi.org/10.1002/hbm.21297

Lee K, Khoo HM, Lina J-M, Dubeau F, Gotman J, Grova C. Disruption, emergence and lateralization of brain network hubs in mesial temporal lobe epilepsy. Neuroimage Clin [Internet]. 2018 Jun 30;20:71–84. Available from: http://dx.doi.org/10.1016/j.nicl.2018.06.029

Lopes da Silva F. EEG and MEG: relevance to neuroscience. Neuron [Internet]. 2013 Dec 4;80(5):1112–28. Available from: http://dx.doi.org/10.1016/j.neuron.2013.10.017

López-Sanz D, Garcés P, Álvarez B, Delgado-Losada ML, López-Higes R, Maestú F. Network Disruption in the Preclinical Stages of Alzheimer’s Disease: From Subjective Cognitive Decline to Mild Cognitive Impairment. Int J Neural Syst [Internet]. 2017 Dec;27(8):1750041. Available from: http://dx.doi.org/10.1142/S0129065717500411

Maris E, Oostenveld R. Nonparametric statistical testing of EEG- and MEG-data. J Neurosci Methods [Internet]. 2007 Aug 15;164(1):177–90. Available from: http://dx.doi.org/10.1016/j.jneumeth.2007.03.024

Marois R, Ivanoff J. Capacity limits of information processing in the brain. Trends Cogn Sci [Internet]. 2005 Jun;9(6):296–305. Available from: http://dx.doi.org/10.1016/j.tics.2005.04.010

Mathewson KE, Gratton G, Fabiani M, Beck DM, Ro T. To see or not to see: prestimulus alpha phase predicts visual awareness. J Neurosci [Internet]. 2009 Mar 4;29(9):2725–32. Available from: http://dx.doi.org/10.1523/JNEUROSCI.3963-08.2009

McCormick C, Quraan M, Cohn M, Valiante TA, McAndrews MP. Default mode network connectivity indicates episodic memory capacity in mesial temporal lobe epilepsy. Epilepsia [Internet]. 2013 May;54(5):809–18. Available from: http://dx.doi.org/10.1111/epi.12098

van Mierlo P, Höller Y, Focke NK, Vulliemoz S. Network Perspectives on Epilepsy Using EEG/MEG Source Connectivity. Front Neurol [Internet]. 2019 Jul 17;10:721. Available from: http://dx.doi.org/10.3389/fneur.2019.00721

van Mierlo P, Papadopoulou M, Carrette E, Boon P, Vandenberghe S, Vonck K, et al. Functional brain connectivity from EEG in epilepsy: seizure prediction and epileptogenic focus localization. Prog Neurobiol [Internet]. 2014 Oct;121:19–35. Available from: http://dx.doi.org/10.1016/j.pneurobio.2014.06.004

Muthukumaraswamy SD, Singh KD. A cautionary note on the interpretation of phase-locking estimates with concurrent changes in power. Clin Neurophysiol [Internet]. 2011 Nov;122(11):2324–5. Available from: http://dx.doi.org/10.1016/j.clinph.2011.04.003

Newman MEJ. The mathematics of networks. The new palgrave encyclopedia of economics [Internet]. 2008;2(2008):1–12. Available from: http://citeseerx.ist.psu.edu/viewdoc/download?doi=10.1.1.131.8175&rep=rep1&type=pdf

Niso G, Carrasco S, Gudín M, Maestú F, Del-Pozo F, Pereda E. What graph theory actually tells us about resting state interictal MEG epileptic activity. Neuroimage Clin [Internet]. 2015 May 23;8:503–15. Available from: http://dx.doi.org/10.1016/j.nicl.2015.05.008

Ofer I, LeRose C, Mast H, LeVan P, Metternich B, Egger K, et al. Association between seizure freedom and default mode network reorganization in patients with unilateral temporal lobe epilepsy. Epilepsy Behav [Internet]. 2019 Jan;90:238–46. Available from: http://dx.doi.org/10.1016/j.yebeh.2018.10.025

Olde Dubbelink KTE, Hillebrand A, Stoffers D, Deijen JB, Twisk JWR, Stam CJ, et al. Disrupted brain network topology in Parkinson’s disease: a longitudinal magnetoencephalography study. Brain [Internet]. 2014 Jan;137(Pt 1):197–207. Available from: http://dx.doi.org/10.1093/brain/awt316

Oostenveld R, Fries P, Maris E, Schoffelen J-M. FieldTrip: Open source software for advanced analysis of MEG, EEG, and invasive electrophysiological data. Comput Intell Neurosci [Internet]. 2011;2011:156869. Available from: http://dx.doi.org/10.1155/2011/156869

Panzica F, Varotto G, Rotondi F, Spreafico R, Franceschetti S. Identification of the Epileptogenic Zone from Stereo-EEG Signals: A Connectivity-Graph Theory Approach. Front Neurol [Internet]. 2013 Nov 6;4:175. Available from: http://dx.doi.org/10.3389/fneur.2013.00175

Papo D, Zanin M, Martínez JH, Buldú JM. Beware of the Small-World Neuroscientist! Front Hum Neurosci [Internet]. 2016 Mar 8;10(96):96. Available from: http://dx.doi.org/10.3389/fnhum.2016.00096

Pedersen M, Omidvarnia AH, Walz JM, Jackson GD. Increased segregation of brain networks in focal epilepsy: An fMRI graph theory finding. Neuroimage Clin [Internet]. 2015 May 22;8:536–42. Available from: http://dx.doi.org/10.1016/j.nicl.2015.05.009

Pion-Tonachini L, Kreutz-Delgado K, Makeig S. ICLabel: An automated electroencephalographic independent component classifier, dataset, and website. Neuroimage [Internet]. 2019 Sep;198:181–97. Available from: http://dx.doi.org/10.1016/j.neuroimage.2019.05.026

Qianqian Fang, Boas DA. Tetrahedral mesh generation from volumetric binary and grayscale images. In: 2009 IEEE International Symposium on Biomedical Imaging: From Nano to Macro [Internet]. 2009. p. 1142–5. Available from: http://dx.doi.org/10.1109/ISBI.2009.5193259

Quraan MA, McCormick C, Cohn M, Valiante TA, McAndrews MP. Altered resting state brain dynamics in temporal lobe epilepsy can be observed in spectral power, functional connectivity and graph theory metrics. PLoS One [Internet]. 2013 Jul 26;8(7):e68609. Available from: http://dx.doi.org/10.1371/journal.pone.0068609

Raghavachari S, Lisman JE, Tully M, Madsen JR, Bromfield EB, Kahana MJ. Theta oscillations in human cortex during a working-memory task: evidence for local generators. J Neurophysiol [Internet]. 2006 Mar;95(3):1630–8. Available from: http://dx.doi.org/10.1152/jn.00409.2005

Ridley BGY, Rousseau C, Wirsich J, Le Troter A, Soulier E, Confort-Gouny S, et al. Nodal approach reveals differential impact of lateralized focal epilepsies on hub reorganization. Neuroimage [Internet]. 2015 Sep;118:39–48. Available from: http://dx.doi.org/10.1016/j.neuroimage.2015.05.096

Rodríguez-Cruces R, Bernhardt BC, Concha L. Multidimensional associations between cognition and connectome organization in temporal lobe epilepsy. Neuroimage [Internet]. 2020 Jun;213:116706. Available from: http://dx.doi.org/10.1016/j.neuroimage.2020.116706

Rosenblum MG, Pikovsky AS, Kurths J. Phase synchronization of chaotic oscillators. Phys Rev Lett [Internet]. 1996 Mar 11;76(11):1804–7. Available from: http://dx.doi.org/10.1103/PhysRevLett.76.1804

Rossini PM, Di Iorio R, Bentivoglio M, Bertini G, Ferreri F, Gerloff C, et al. Methods for analysis of brain connectivity: An IFCN-sponsored review. Clin Neurophysiol [Internet]. 2019 Oct;130(10):1833–58. Available from: http://dx.doi.org/10.1016/j.clinph.2019.06.006

Rubinov M, Sporns O. Complex network measures of brain connectivity: uses and interpretations. Neuroimage [Internet]. 2010 Sep;52(3):1059–69. Available from: http://dx.doi.org/10.1016/j.neuroimage.2009.10.003

Rubinov M, Sporns O. Weight-conserving characterization of complex functional brain networks. Neuroimage [Internet]. 2011 Jun 15;56(4):2068–79. Available from: http://dx.doi.org/10.1016/j.neuroimage.2011.03.069

Sauseng P, Klimesch W, Heise KF, Gruber WR, Holz E, Karim AA, et al. Brain oscillatory substrates of visual short-term memory capacity. Curr Biol [Internet]. 2009 Nov 17;19(21):1846–52. Available from: http://dx.doi.org/10.1016/j.cub.2009.08.062

Schoffelen J-M, Gross J. Source connectivity analysis with MEG and EEG. Hum Brain Mapp [Internet]. 2009 Jun;30(6):1857–65. Available from: http://dx.doi.org/10.1002/hbm.20745

Shine JM, Breakspear M, Bell PT, Ehgoetz Martens KA, Shine R, Koyejo O, et al. Human cognition involves the dynamic integration of neural activity and neuromodulatory systems. Nat Neurosci [Internet]. 2019 Feb;22(2):289–96. Available from: http://dx.doi.org/10.1038/s41593-018-0312-0

Sion A, Bruña Fernández R, Martínez Maldonado A, Domínguez Centeno I, Torrado-Carvajal A, Rubio G, et al. Resting-state connectivity and network parameter analysis in alcohol-dependent males. A simultaneous EEG-MEG study. J Neurosci Res [Internet]. 2020 Jun 25; Available from: http://dx.doi.org/10.1002/jnr.24673

Staljanssens W, Strobbe G, Van Holen R, Keereman V, Gadeyne S, Carrette E, et al. EEG source connectivity to localize the seizure onset zone in patients with drug resistant epilepsy. Neuroimage Clin [Internet]. 2017 Sep 14;16:689–98. Available from: http://dx.doi.org/10.1016/j.nicl.2017.09.011

Stam CJ. Modern network science of neurological disorders. Nat Rev Neurosci [Internet]. 2014 Oct;15(10):683–95. Available from: http://dx.doi.org/10.1038/nrn3801

Stam CJ, Tewarie P, Van Dellen E, van Straaten ECW, Hillebrand A, Van Mieghem P. The trees and the forest: Characterization of complex brain networks with minimum spanning trees. Int J Psychophysiol [Internet]. 2014 Jun;92(3):129–38. Available from: http://dx.doi.org/10.1016/j.ijpsycho.2014.04.001

Stefan H, Lopes da Silva FH. Epileptic neuronal networks: methods of identification and clinical relevance. Front Neurol [Internet]. 2013 Mar 1;4:8. Available from: http://dx.doi.org/10.3389/fneur.2013.00008

van Straaten ECW, Stam CJ. Structure out of chaos: functional brain network analysis with EEG, MEG, and functional MRI. Eur Neuropsychopharmacol [Internet]. 2013 Jan;23(1):7–18. Available from: http://dx.doi.org/10.1016/j.euroneuro.2012.10.010

Tailby C, Kowalczyk MA, Jackson GD. Cognitive impairment in epilepsy: the role of reduced network flexibility. Ann Clin Transl Neurol [Internet]. 2018 Jan;5(1):29–40. Available from: http://dx.doi.org/10.1002/acn3.503

Tewarie P, van Dellen E, Hillebrand A, Stam CJ. The minimum spanning tree: an unbiased method for brain network analysis. Neuroimage [Internet]. 2015 Jan 1;104:177–88. Available from: http://dx.doi.org/10.1016/j.neuroimage.2014.10.015

Tzourio-Mazoyer N, Landeau B, Papathanassiou D, Crivello F, Etard O, Delcroix N, et al. Automated anatomical labeling of activations in SPM using a macroscopic anatomical parcellation of the MNI MRI single-subject brain. Neuroimage [Internet]. 2002 Jan;15(1):273–89. Available from: http://dx.doi.org/10.1006/nimg.2001.0978

Uhlhaas PJ, Singer W. Neural synchrony in brain disorders: relevance for cognitive dysfunctions and pathophysiology. Neuron [Internet]. 2006 Oct 5;52(1):155–68. Available from: http://dx.doi.org/10.1016/j.neuron.2006.09.020

Van Veen BD, van Drongelen W, Yuchtman M, Suzuki A. Localization of brain electrical activity via linearly constrained minimum variance spatial filtering. IEEE Trans Biomed Eng [Internet]. 1997 Sep;44(9):867–80. Available from: http://dx.doi.org/10.1109/10.623056

Vaughan DN, Rayner G, Tailby C, Jackson GD. MRI-negative temporal lobe epilepsy: A network disorder of neocortical connectivity. Neurology [Internet]. 2016 Nov 1;87(18):1934–42. Available from: http://dx.doi.org/10.1212/WNL.0000000000003289

Vecchio F, Miraglia F, Curcio G, Della Marca G, Vollono C, Mazzucchi E, et al. Cortical connectivity in fronto-temporal focal epilepsy from EEG analysis: A study via graph theory. Clin Neurophysiol [Internet]. 2015 Jun;126(6):1108–16. Available from: http://dx.doi.org/10.1016/j.clinph.2014.09.019

Vecchio F, Miraglia F, Quaranta D, Granata G, Romanello R, Marra C, et al. Cortical connectivity and memory performance in cognitive decline: A study via graph theory from EEG data. Neuroscience [Internet]. 2016 Mar 1;316:143–50. Available from: http://dx.doi.org/10.1016/j.neuroscience.2015.12.036

van Veenendaal TM, Ijff DM, Aldenkamp AP, Lazeron RHC, Hofman PAM, de Louw AJA, et al. Chronic antiepileptic drug use and functional network efficiency: A functional magnetic resonance imaging study. World J Radiol [Internet]. 2017 Jun 28;9(6):287–94. Available from: http://dx.doi.org/10.4329/wjr.v9.i6.287

Vlooswijk MCG, Vaessen MJ, Jansen JFA, de Krom MCFTM, Majoie HJM, Hofman PAM, et al. Loss of network efficiency associated with cognitive decline in chronic epilepsy. Neurology [Internet]. 2011 Sep 6;77(10):938–44. Available from: http://dx.doi.org/10.1212/WNL.0b013e31822cfc2f

Vorwerk J, Cho J-H, Rampp S, Hamer H, Knösche TR, Wolters CH. A guideline for head volume conductor modeling in EEG and MEG. Neuroimage [Internet]. 2014 Oct 15;100:590–607. Available from: http://dx.doi.org/10.1016/j.neuroimage.2014.06.040

Výtvarová E, Mareček R, Fousek J, Strýček O, Rektor I. Large-scale cortico-subcortical functional networks in focal epilepsies: The role of the basal ganglia. Neuroimage Clin [Internet]. 2017;14:28–36. Available from: http://dx.doi.org/10.1016/j.nicl.2016.12.014

Watts DJ, Strogatz SH. Collective dynamics of “small-world” networks. Nature [Internet]. 1998 Jun 4;393(6684):440–2. Available from: http://dx.doi.org/10.1038/30918

van Wijk BCM, Stam CJ, Daffertshofer A. Comparing brain networks of different size and connectivity density using graph theory. PLoS One [Internet]. 2010 Oct 28;5(10):e13701. Available from: http://dx.doi.org/10.1371/journal.pone.0013701

